# Barriers in recipient partners cause frequent transient HIV infections and explain transmission risk under viral sup­pression

**DOI:** 10.64898/2026.03.20.26348904

**Authors:** Katherine E. Atkins, Tibor Antal, Robin N. Thompson, Katrina Lythgoe, Roland Regoes, Stéphane Hué, Ch. Julián Villabona-Arenas

## Abstract

Treatment-as-prevention is the cornerstone of global HIV control, and the ‘Undetectable = Untransmittable’ (U=U) message rests on the empirical observation that antiretroviral therapy reduces onward transmission to negligible levels. Yet a mechanistic explanation for why viral suppression prevents transmission, and for the broader quantitative features of HIV transmission, has been lacking. Here we develop a mechanistic framework that resolves three long-standing features simultaneously: the low per-act transmission probability, the narrowing of within-host viral diversity to one or a handful of founder variants, and the plateau of transmission risk at high viral loads. We show that three biologically grounded mechanisms – short windows of susceptibility in the exposed partner, stage-dependent establishment of systemic infection, and target-cell limitation at the site of infection – are jointly necessary and sufficient to reconcile these observations. Calibrated to six epidemiological datasets and independently validated against three more, the model derives a transmission rate below 0.05 systemic infections per 100 couple-years follow-up under viral suppression, providing the mechanistic basis for the negligible risk observed in PARTNER1 and the U=U message that rests on it. Recalibration to data from men who have sex with men attributes their elevated transmission rates to longer windows of susceptibility, consistent with the biology of the rectal mucosa, rather than to differences in per-act biology – reframing how population-level risk differences should be interpreted in prevention planning. The model further predicts four to five transient infections for each systemic infection, consistent with evidence from the STEP vaccine trial and independently supported by HIV-specific immune responses in highly exposed seronegative individuals and transient local viral RNA in tissues from non-human primate challenge studies. Embedding the model in a phylodynamic framework substantially narrows estimates of when transmission occurred and how many viral variants established infection. Together, these findings provide a mechanistic foundation for treatment-as-prevention, reframe the determinants of population-level transmission differences, and point to vaccine strategies aimed at preventing systemic establishment after mucosal exposure.

## Introduction

Empirical data suggest a complex and seemingly contradictory picture of HIV transmission. While untreated individuals can experience high levels of viral load in semen or cervicovaginal compartments [1–3], the estimated probability of HIV acquisition per sexual exposure remains low; for example, for heterosexual couples, this risk is estimated as approximately 0.18% per act [4]. These observations suggest that stringent transmission barriers must limit viral transmission during sexual exposure [5]. However, HIV infections are often initiated by multiple genetic variants [6], suggesting that any transmission barriers can be overcome by multiple virions simultaneously.

One potential explanation for this paradox is that conditions permissive to HIV infection arise only intermittently, e.g. when the genital epithelial barrier is compromised by microabrasions or when inflammatory co-infections are present. When such permissive conditions occur, genetic bottlenecks are less stringent; for example, genital ulcers increase the estimated per-act transmission probability by approximately 5.3 times [4]. This ‘intermittent susceptibility’ model of HIV transmission can reconcile the apparent contradiction between stringent barriers and multi-variant transmission [7]. However, one observation remains unexplained by this model: that transmission risk becomes uncoupled from viral load at high set-point viral loads [8–12]. In addition, although the intermittent susceptibility model assumes that the probability of HIV acquisition varies across infection stages—being higher during early and late stages than during the asymptomatic stage [4, 13]—these observations were not matched against empirical stage-specific risks. A mechanistic model that can capture all of these epidemiological observations is therefore currently unavailable.

While the intermittent susceptibility model is supported by empirical evidence suggesting that biological barriers within the exposed partner reduce the risk of transmission [14, 15], two additional mechanisms may also shape transmission outcomes. First, viral infectivity in the transmitting partner may vary over the course of infection due to within-host viral evolution under immune pressure [16–19]. Second, shortly after exposure, stochastic effects in the exposed partner—including target cell availability [20–22] and local immune responses [23–26]—may determine whether systemic infection is established and which viral lineages succeed [5, 27–30]. Whether these mechanisms leave a detectable imprint on population-level HIV epidemiology has not been tested.

Here we show that three biologically grounded mechanisms—intermittent susceptibility, stage-dependent infectivity, and target cell limitation—are sufficient to reconcile key features of HIV transmission. We developed seven transmission models encoding different combinations of these mechanisms and calibrated each against population-level epidemiological data. We then validated our best-fitting model against phylogenetic and epidemiological data from 48 individual transmission pairs. Our best-fitting model reveals that transient infections are substantially more common than systemic infections, provides a mechanistic explanation for the negligible transmission risk under viral suppression, and shows that differences in transmission rates between heterosexual and MSM populations are primarily explained by a greater probability of permissive conditions for infection—predictions we further validated against large-scale HIV studies.

## Results

To understand the mechanisms governing the transmission of HIV during sexual exposure, we constructed seven mechanistic models of viral transmission (M1-M7), each encoding a different combination of intermittent susceptibility, stage-dependent infectivity, and target-cell limitation. To choose the most parsimonious model that was able to capture empirical HIV data, we fit each model separately using Markov chain Monte Carlo to six population-level observations of heterosexual HIV transmission: (i) the per-act acquisition probability during the asymptomatic stage; (ii) the hazards of acquisition during the early and late stages relative to the asymptomatic stage; (iv) the per-act probability of multiple-variant transmission; (v) the relative hazard of multiple founder variants during the early versus asymptomatic stage; and (vi) the annual transmission rate stratified by set-point viral load. Full model derivations, priors, and the fitting procedure are given in Methods.

### Transmission Model Calibration

Our analysis revealed that model M7—which encodes intermittent susceptibility, stage-dependent virus infectivity, and target-cell limitation—best and most parsimoniously captured the six key empirical observations of HIV transmission dynamics (**Figure 1**, **Supplementary Figure 1**). First, the model captures the stage-specific risks observed in the data by assigning stage-specific differences to per-infected-cell establishment probabilities rather than per-virion acquisition probabilities (**Supplementary Figure 1**), with the mean per-cell probability of establishment during the early stage of infection (*r_E_* = 0.28) being substantially higher than that of the late stage (*r_L_* = 0.01), which was in turn higher than that of the asymptomatic stage (*r_A_*= 0.003) (**Figure 2**, **Supplementary Table 1**). Second, the model captures the low probability of acquisition but a high frequency of multiple-variant infection by estimating a low fraction of exposures permissive to infection (*f* = 0.7%, 95% HPD 0.4–1.0%). Third, the model captures the plateauing transmission rate with increasing viral loads through target cell limitation, with a posterior mean of 300 target cells at the exposure site (95% HPD 6-861), such that there was no increase in transmission rate once the set-point viral load had reached 4.5 log copies/mL.

**Figure 1:**
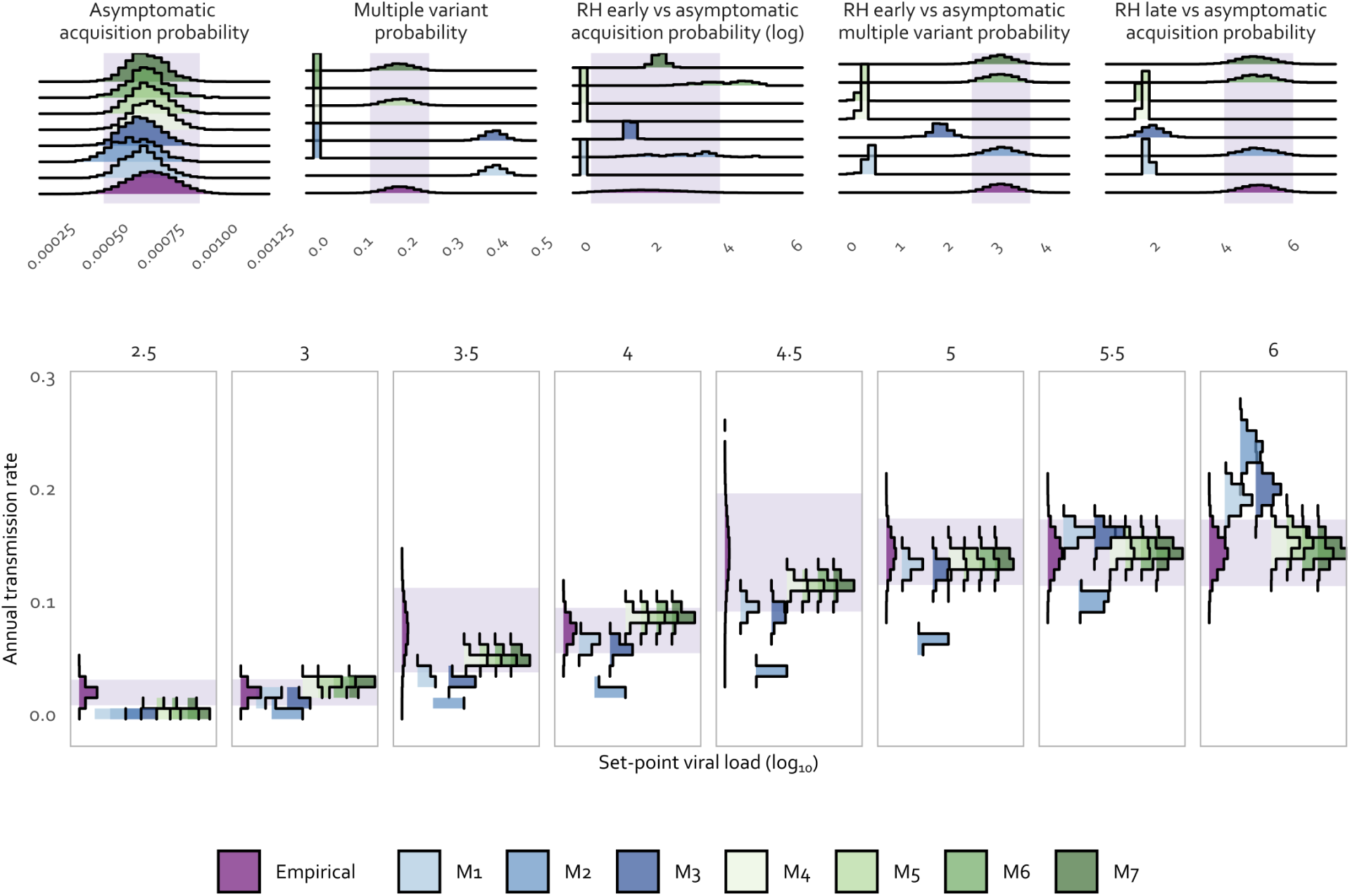
Model M7 best captures all empirical observations of heterosexual HIV transmission. Top panels display model fitting results for five empirical epidemiological metrics: asymptomatic acquisition probability, multiple variant probability, relative hazard (RH) comparisons between early vs. asymptomatic acquisition (shown in log scale), RH of early vs. asymptomatic multiple variant probability, and RH of late vs. asymptomatic acquisition probability. Bottom panels show model fitting results for annual transmission rates across different viral load set points (3.3 to 5.8 in log_10_ scale), plotted as rotated figures to evidence how annual transmission rates plateau as viral load increases. Purple distributions represent the empirical data being fitted with purple bars indicating the 2.5th and 97.5th percentile range. Seven distinct models are compared using different colors: M1 (Intermittent susceptibility), M2 (Stage-dependent virus infectivity), M3 (Intermittent susceptibility, stage-dependent virus infectivity), M4 (target-cell-limited), M5 (Intermittent susceptibility & target-cell-limited), M6 (stage-dependent virus infectivity & target-cell-limited), and M7 (Intermittent susceptibility, stage-dependent virus infectivity and target-cell-limited).

**Figure 2:**
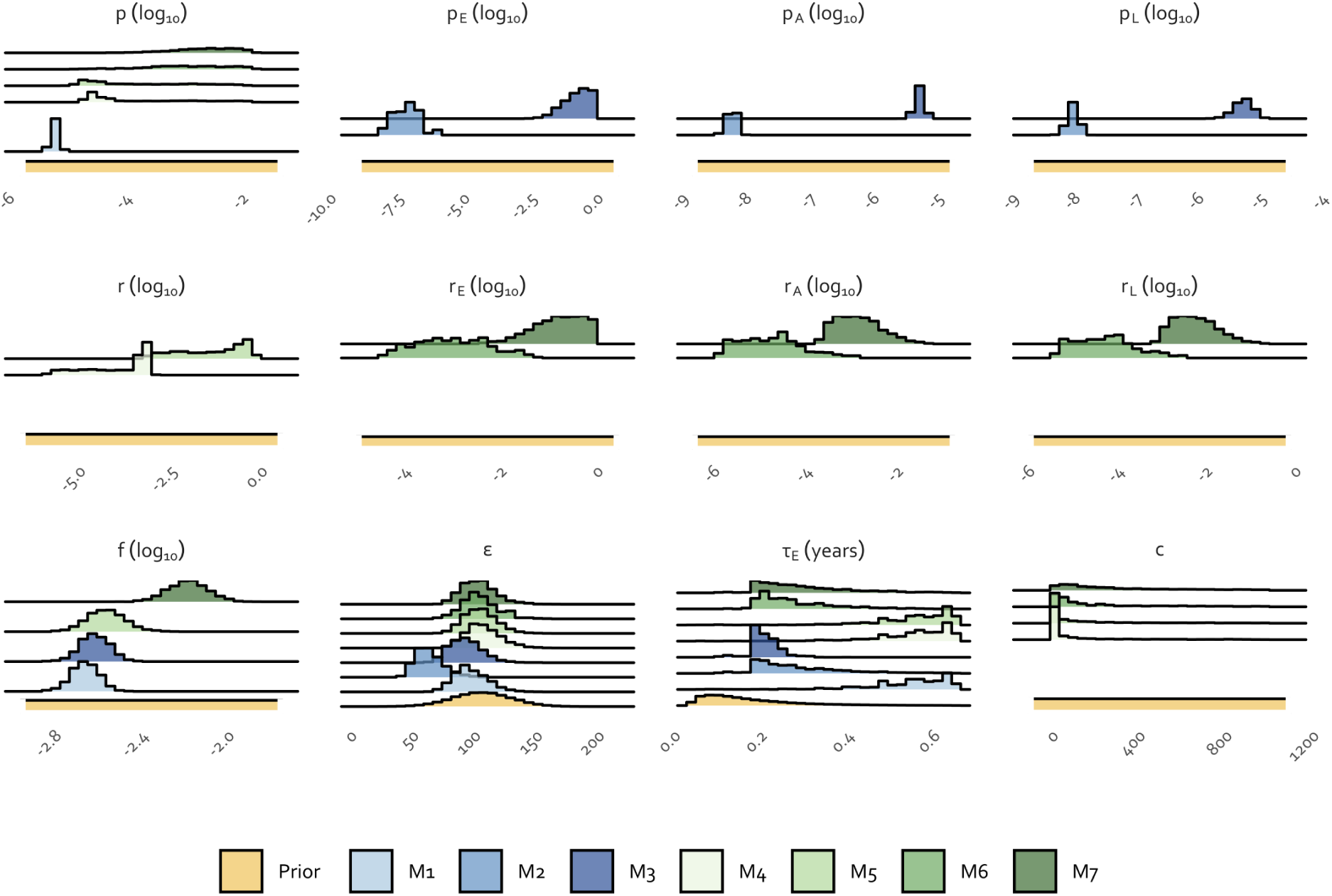
Posterior parameter estimates vary across models, reflecting different mechanistic explanations of the same data. *p*, Per-virion acquisition probability (*p_E_*, *p_A_* and *p_L_* for early, asymptomatic and late stages, respectively); *r*, per-infected-cell establishment probability (*r_E_*, *r_A_* and *r_L_* for early, asymptomatic and late stages, respectively); *f*, probability of permissive conditions for infection; *ε*, number of sexual exposures per year between an uninfected and an asymptomatically infected individual; *τ_E_*, duration of the early stage of infection in years; *c*, number of target cells available for initial infection in the exposed partner. Absent distributions in individual panels indicate that the corresponding parameter is not included in that particular model formulation. Seven distinct models are compared using different colors: M1 (Intermittent susceptibility), M2 (Stage-dependent virus infectivity), M3 (Intermittent susceptibility, stage-dependent virus infectivity), M4 (target-cell-limited), M5 (Intermittent susceptibility & target-cell-limited), M6 (stage-dependent virus infectivity & target-cell-limited), and M7 (Intermittent susceptibility, stage-dependent virus infectivity and target-cell-limited).

The correlated posterior distributions suggest that while the model’s predictions are well constrained by the fitted observations, the relative contributions of cell infection versus establishment to the transmission bottleneck are less identifiable individually (**Supplementary Figure 2**). For example, the number of cells (*c*) and the per-virion acquisition probability (*p*) were positively correlated, as were the three establishment probabilities (*r_E_*, *r_A_*, *r_L_*), but these two groups were negatively correlated.

When we recalibrated our model to men who have sex with men (MSM) transmission, the probability of permissive conditions for infection was approximately 2.5 times (from 0.7% to 1.7%; 95% HPD 0.9–2.5%) that of the heterosexual population (**Supplementary Table 1, Supplementary Figure 3**). Consistent with this finding, the mean number of sexual exposures per year—to achieve the maximum transmission risk—decreased by more than half, from 104 (95% HPD 77–129) to 49 (95% HPD 31–69).

In sensitivity analyses, excluding RH_Mult_ from the fitted observations resulted in a shorter estimated duration of the early stage (*τ_E_*decreased from 110 days to 66 days, 95% HPD 15–150). Credible intervals for the remaining parameters overlapped broadly between the two analyses, with substantially wider intervals for *r_A_* and *r_L_* when RH_Mult_ was excluded, suggesting that this observation helps constrain the stage-specific establishment probabilities (**Supplementary Table 1, Supplementary Figure 4**).

### Individual-level phylodynamic analysis

We embedded our best-fitting transmission model within a coalescent-based phylodynamic framework to estimate the time since infection (TSI) and viral load at transmission for 48 individual transmission pairs. Preliminary analyses showed that phylogenetic summary statistics better distinguished between one versus two founding variants when the time since infection was longer (**Supplementary Figure 5**; see **Supplementary Results** for details). The Approximate Bayesian Computation Sequential Monte Carlo (ABC-SMC) fitting procedure produced good overall agreement between simulated and empirical phylogenies (**Supplementary Figures 6, 7**). Phylogenetic data informed transmission timing parameters, with posterior distributions of time since infection narrowing relative to priors in 27 of 43 cases (63%) (narrowing defined as an interquartile range ratio below 0.50; **Figure 3**). Likewise, viral load posteriors narrowed relative to priors in 6 of 8 early transmitter scenarios (**Supplementary Figure 8**).

**Figure 3:**
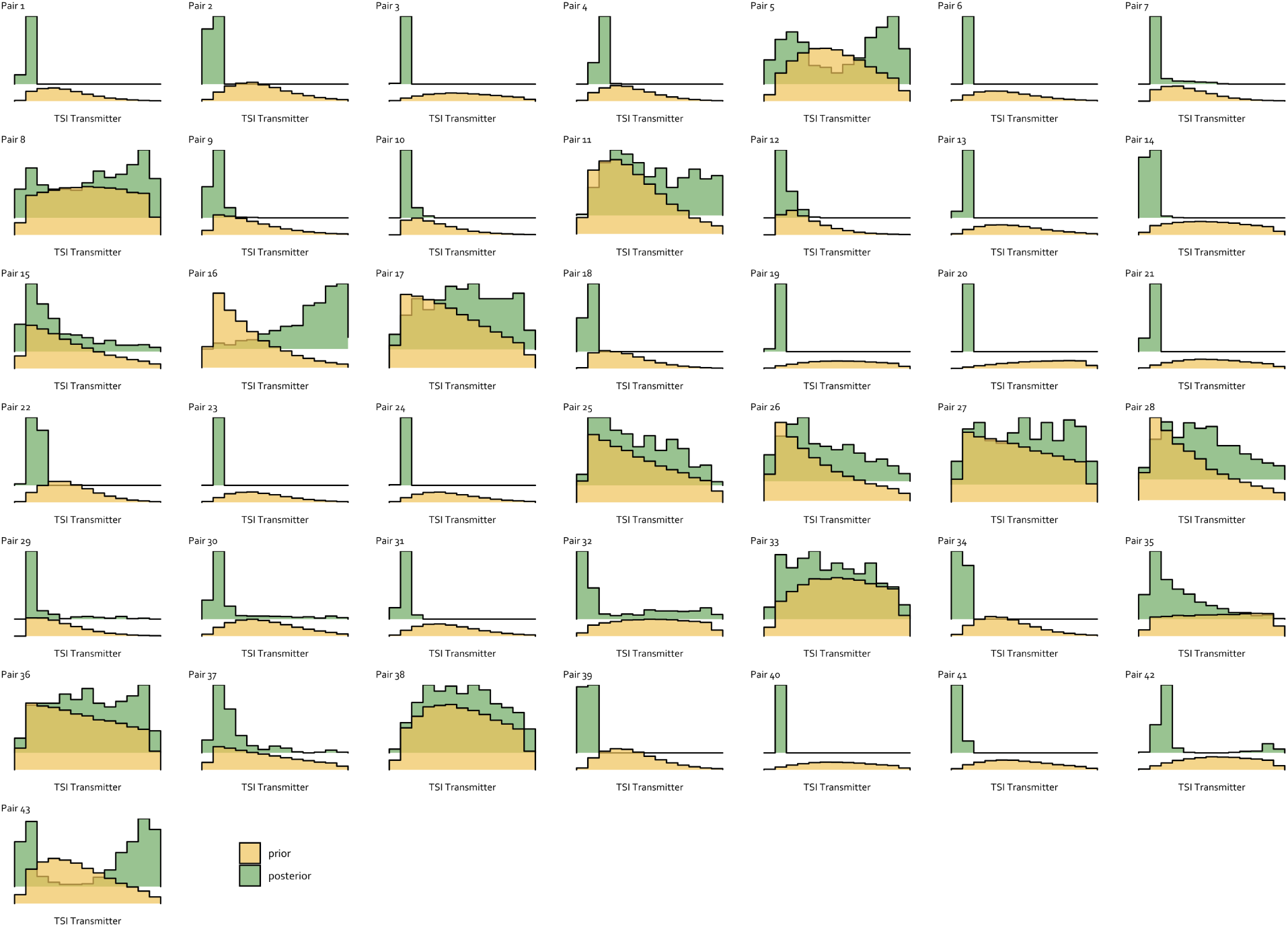
Phylogenetic data narrow posterior estimates of time since infection for the majority of transmission pairs. Each panel represents one transmission pair. Prior distributions reflect an epidemiologically plausible range of time since infection, and posterior distributions incorporate phylogenetic data through an ABC-SMC fitting procedure.

### Virions and variants

Using the best-fitting calibrated model, we characterised the relationship between transmitted virions and the number of genetically distinct viral lineages (founder variants) establishing infection across infection stages (**Figure 4, Supplementary Table 2**). Early-stage infections were limited by low viral diversity: high viral loads and high per-cell establishment probability enabled multiple virion transmission (89%), but low diversity meant that most transmissions involved a single variant (68%) or multiple variants (21%). Despite high virion numbers, the number of founding variants during early-stage transmissions rarely exceeded two (**Figure 4A**). Asymptomatic infections were limited by low viral counts, with transmission of mostly a single virion and variant (91%). When multiple virions were transmitted, high diversity enabled more multiple founder variants (7%) than single variants (2%). In late-stage transmissions, founding virions and variants were more correlated than in early-stage: when multiple virions were transmitted, high diversity meant that these were more likely to represent distinct variants (34% multiple variants). However, the majority of late-stage transmissions still involved a single virion and variant (63%). Stratifying these results by the transmitting partner’s viral load showed that high viral loads increased the number of founding virions across all stages, but only translated into more founding variants during late-stage infections where viral diversity was high (**Figure 4B**).

**Figure 4:**
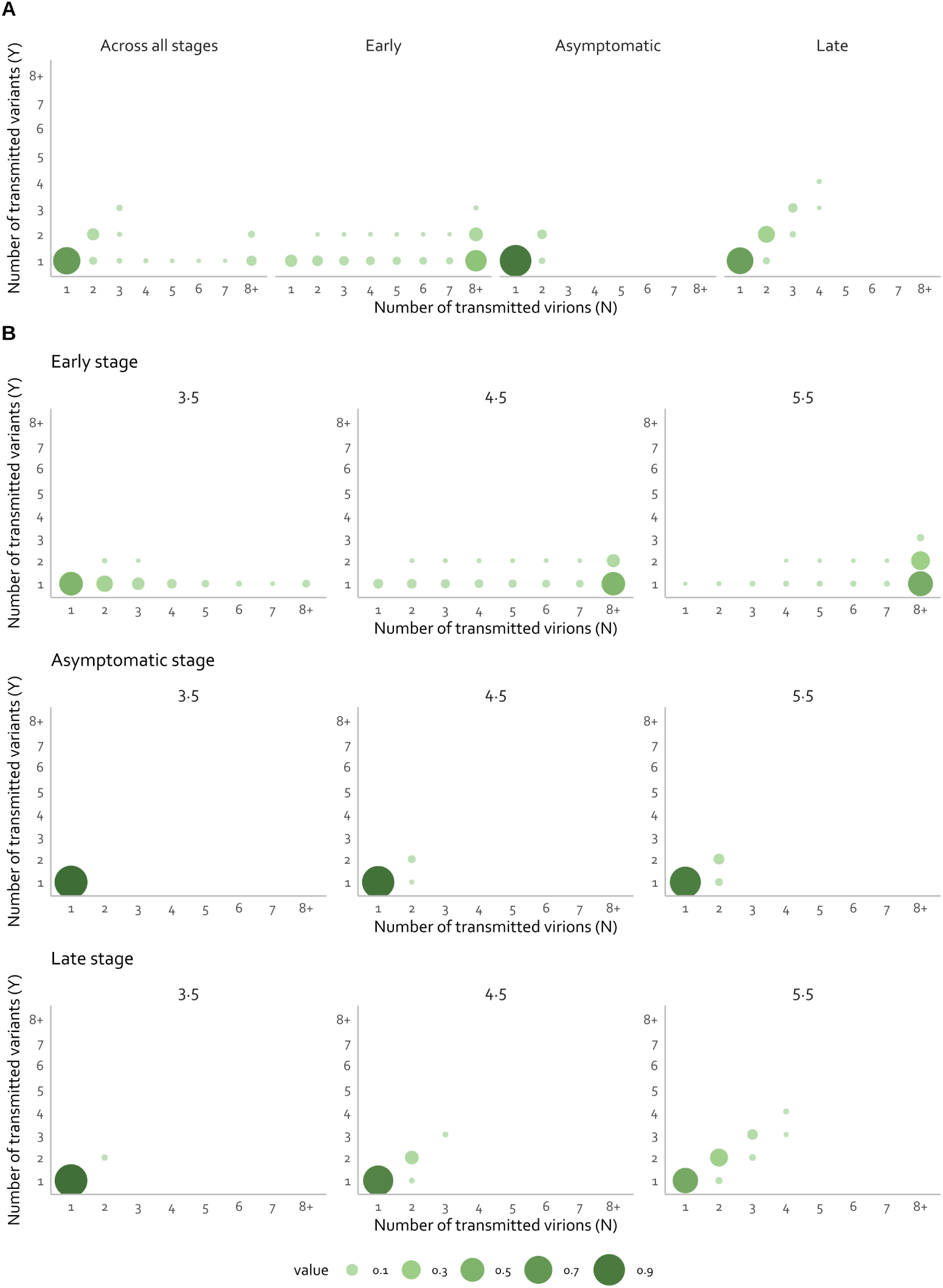
The number of founding variants is closely linked to transmitted virions but modulated by infection stage. Each panel shows the joint probability of the effective number of transmitted virions and variants establishing infection in the exposed partner either by stage (column)**(A)**, or by stage (rows) for a given log_10_ viral load (columns) **(B)**.

When recalibrated to MSM transmission—which was fitted to a higher probability of multiple variant transmission (∼30% versus ∼19% for heterosexuals)—the proportion of multiple variant transmission increased across all stages, particularly during the late stage where multiple variant transmission became the majority (58% versus 34% in heterosexuals) (**Supplementary Table 2**).

In the sensitivity analysis excluding the relative hazard of multiple founder variants during the early versus asymptomatic stage (RH_Mult_) from the fitted observations, the model predicted a RH_Mult_ of approximately 0.95. Consistent with this observation, the early stage risk shifted substantially, with single virion and variant transmissions increasing from 11% to 46% and multiple variant transmission dropping from 21% to 4%. However, the overall proportion of multiple variant transmission across all stages remained at 18%, as the reduced early-stage contribution was offset by a greater relative contribution from late-stage transmissions, where multiplicity was largely unchanged (**Supplementary Table 2**).

### Transmission risk, cell infection, and transient infections

After fitting our model to empirical stage-specific risks, we predicted an overall per-act transmission risk of 0.12% (95% HPD 0.09–0.15%) (**Figure 5A**), consistent with the lower end of the empirical estimate of 0.18% (95% CI 0.11–0.30%) reported by [4].

**Figure 5:**
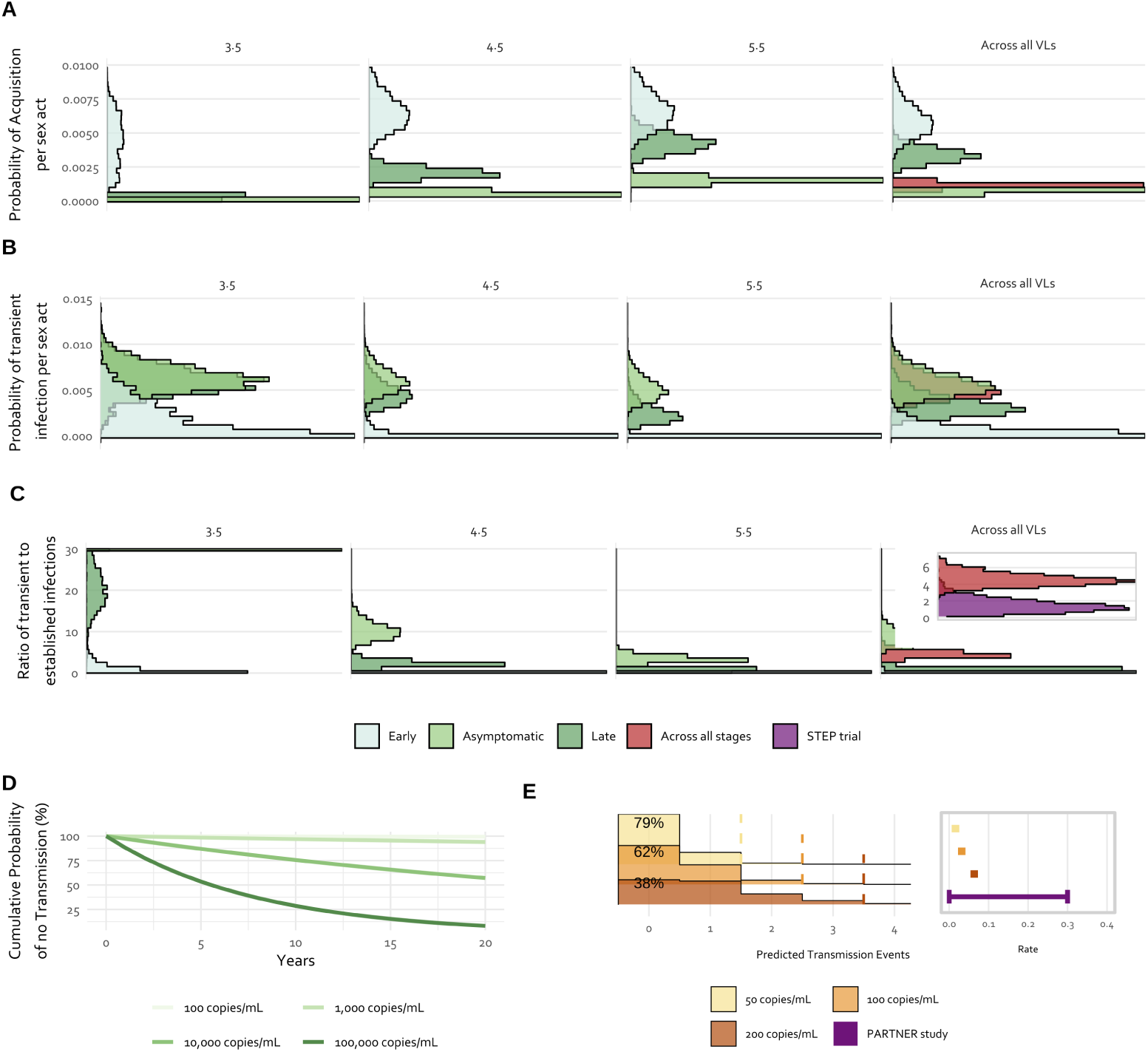
The calibrated model predicts low transmission risk, frequent transient infections, and large risk reduction under viral suppression. **(A)** Probability of acquisition per sexual act by stage of infection (colours) and by a given viral load (numbers above each panel). **(B)** Fraction of failed transmission attempts that result in transient infections. **(C)** Ratio of transient to established infections. For log_10_ VL = 3.5 panel, density mass exceeding the axis limit is compressed to the boundary. The rightmost panel includes a zoomed inset comparing the ‘Across all stages’ prediction to the minimum ratio observed in the STEP trial. **(D)** Cumulative probability of no transmission during the asymptomatic stage. **(E)** Transmission events based on the PARTNER1 study population characteristics (1166 couples, 48 exposures per couple over 1.3 years of follow-up). The dashed lines represent the upper bound of the 95% confidence intervals. The right inset shows the predicted rate of within-couple per 100 years-couple of follow up and the 95% confidence range found in the PARTNER1 study.

Our model predicts that the expected proportion of target cells at the exposure site that become infected, under permissive conditions, is around 41% (95% HPD 34–46%) for the early and asymptomatic stages, and around 69% for the late stage (95% HPD 59–75%). Given that cells become infected, the median probability of transient infection—the chance that infection is stochastically cleared—is 0.82 (95% HPD 0.77–0.86) (**Supplementary Table 3**). The probability of transient infection varies across infection stage, with the highest probability for the asymptomatic stage (0.89; 95% HPD 0.85–0.92), followed by the late stage (0.49; 95% HPD 0.35–0.64) and the early stage (0.09; 95% HPD 0.002–0.32). Consequently, transient infections occur in 0.53% (95% HPD 0.33–0.82%) of all sexual exposure events across the infection cycle (**Figure 5B**). However, because overall infection risk is low, our model suggests that around 4.6 (95% HPD 3.3–6.3) transient infections occur for each systemic infection (**Figure 5C**), about 3.5 times the lower bound of around 1.3 suggested by empirical data [31].

In sensitivity analyses, the overall model predictions remained consistent when excluding RH_Mult_ from the fitting procedure (**Supplementary Table 3**). Recalibrating our model to MSM transmission increased the predicted overall transmission risk more than three times (from 0.12% to 0.40%, 95% HPD 0.25–0.56%), while the ratio of transient to established infections decreased from 4.6 to 3.1 (95% HPD 2.3–4.2) indicating that a larger proportion of transmission events in MSM result in systemic infection (**Supplementary Table 3**).

### Transmission risk from virally-suppressed individuals

We used our model to calculate the cumulative risk of transmission during the asymptomatic stage using the median number of condomless sex acts per year (37) reported in the PARTNER1 study [32]. Our results suggest that, with undetectable viral loads, the cumulative risk is very low. For example, a viral load of 50 copies/mL results in approximately 0.02%, 0.08%, and 0.32% cumulative probability of transmission after 1, 5, and 20 years, respectively. Conversely, these risks for untreated individuals with a set-point viral load of 4.5 log copies/mL (∼31,600 copies/mL) are 7%, 29%, and 75% (**Figure 5D**).

We then used our best-fit model to predict the number of transmission events that would have been observed through the PARTNER1 study cohort (1,166 couples with a median follow-up of 1.3 years). Specifically, for a given undetectable viral load, we computed the probability of transmission per couple during the trial follow-up period (1.3y), with the average annual number of unprotected sexual exposures reported in the trial (37), and a per-exposure acquisition probability provided by the model. Treating each couple as an independent Bernoulli trial, the total number of transmissions across the trial follow up follows a binomial distribution. Our model predicts that for individuals with a viral load of 50 copies/mL, the probability of observing zero transmission events is approximately 0.79 (95% prediction interval: 0–1 transmission events across the cohort), with an expected transmission rate of 0.02 per 100 couple-years of follow-up (**Figure 5E**). This aligns closely with the zero transmissions observed in the PARTNER1 study, which reported an upper 95% confidence limit of 0.30 per 100 couple-years of follow-up among virally suppressed individuals (viral load less than 200 copies/mL).

## Discussion

Our study addresses a critical challenge in HIV epidemiology: understanding how characteristics of both the transmitting and exposed partners jointly shape transmission outcomes. We achieved this by using a mathematical model of HIV transmission to reconcile population-level epidemiological observations. First, the contrast between high viral loads and low acquisition probabilities was explained through a low per-virion acquisition probability combined with infrequent permissive conditions for infection, likely reflecting stringent transmission barriers that fluctuate through time. Second, the relatively high probability of multiple founder variants was attributed to relaxed bottlenecks during these rare permissive exposures allowing multiple virions to establish infection simultaneously. Third, the plateauing of transmission rates at high viral loads was explained by target cell limitation at the infection site. Fourth, stage-specific differences in both transmission probability and the frequency of multiple founder variants were attributed to variations in the probability that infected cells establish systemic infection. Beyond reconciling these observations, the calibrated model yields novel predictions: transient infections occur approximately five times more frequently than systemic infections, viral suppression reduces transmission risk by several hundredfold, and higher transmission rates in MSM populations are primarily explained by a greater probability of permissive conditions for infection, that typically occur infrequently.

To our knowledge, this is the first model to reconcile all these key epidemiological observations, and it yields additional insights into HIV transmission. The model predicts a close association between a transmitter’s viral load and the number of virions infecting exposed partners, with the transmitter’s infection stage influencing the number of transmitted variants. These patterns are broadly consistent with previous theoretical predictions [7], with two notable differences: first, our model permits a wider range of particle counts to cross the transmission barrier, reflecting a trade-off between particle counts and the per-cell establishment probability; second, during early infection, our model predicts a higher frequency of two to three founding variants [7], a consequence of fitting to the elevated relative hazard of multiple founders observed at this stage.

These findings offer a mechanistic interpretation of stage-specific transmission risk. The probability that an infected cell establishes systemic infection peaks during early infection, aligning with clinical observations that transmission risk during this stage exceeds what would be expected from elevated viral load alone [33]. This elevated establishment probability may reflect the transmission of highly infectious founder-like variants that are subsequently lost through immune-driven evolution and antibody neutralisation [34–36]. Consistent with this, transmitters in the asymptomatic stage showed markedly lower per-cell establishment probability (which increases again during late infection, and the biological basis for this pattern remains unclear). Alternatively, the stage-dependent variation in establishment probability may be linked to the accumulation of costly immune escape mutations during infection and their impact on viral fitness in new hosts [37, 38]. Host heterogeneity, including HLA type [38], may further modulate these differences.

Our prediction that transient infections occur 4.6 as often as systemic infections aligns with indirect evidence from the STEP trial, which suggests that for every established infection, there may have been at least 1.3 unobserved transient infections [31, 39]. Additional supporting evidence that transient infections occur come from multiple sources: a prospective study of 47 seronegative HIV-exposed individuals found that eight tested positive for early serum HIV antibodies but did not progress to detectable infection, compared with two who became permanently infected [40]; a cohort of highly exposed seronegative female sex workers showed detectable HIV-specific IgA in cervical tissue without systemic seroconversion, with a positive correlation between reported number of sexual contact and presence of HIV-specific IgA [41]; and SIV–macaque models have demonstrated ‘occult’ infections in which provirus (integrated virus) becomes subsequently undetectable [42–44]. Mucosal antibody findings suggest that localised immune responses may reflect transient infection and subsequent stochastic clearance, which is corroborated by modelling of SIV repeat low-dose challenge studies [45].

For virally suppressed individuals, the model predicts very low transmission risk compared with unsuppressed individuals, even with regular sexual exposure to long-term partners. While absolute risk estimates should be interpreted cautiously, the model predicts a several-hundred-fold reduction in cumulative transmission probability for individuals with undetectable viral loads (50 copies/mL). This prediction, while consistent with the strong support for the ‘undetectable equals untransmittable’ (U=U) principle, provides a mechanistic basis for understanding how this transmission risk is reduced.

These mechanistic insights provide theoretical support for several prevention strategies. The high probability that infected cells progress to systemic infection during early infection underscores the critical importance of rapid identification and treatment of newly infected individuals—interventions that could interrupt transmission during the period of greatest per-exposure risk. Similarly, the elevated establishment probability during late-stage infection relative to the asymptomatic stage emphasises the value of regular testing to maintain viral suppression and prevent onward transmission. The model’s emphasis on permissive conditions for successful transmission provides theoretical support for managing co-infections that compromise mucosal barriers.

Our model suggested that higher transmission rates in MSM populations likely result from more chance of permissive conditions for infection. This result aligns with empirical observations: MSM engaging in condomless receptive anal intercourse show distinct rectal mucosa phenotypes compared to men never engaged in anal intercourse, characterised by increased immune activation (more CD8+ T cell proliferation), inflammatory responses (molecular signatures associated with mucosal injury and repair), and changes in microbial composition [14]. All of these characteristics are mechanistically linked to a higher chance of HIV acquisition [46, 47]. Under these more permissive conditions, fewer annual sexual exposures are required for infection in MSM compared to heterosexual individuals.

Our phylodynamic analysis using linked transmission pairs provided preliminary evidence that our transmission model can capture individual transmission dynamics. Transmission pair data offer valuable insights into HIV transmission, but inherent uncertainties in transmission timing and viral measurements complicate their interpretation. We successfully matched simulated data to empirical data through ABC-SMC, demonstrating that our mechanistic model can help reproduce phylogenetic patterns. Our results underscore that phylogenetic inference of transmission history is affected by the duration of infection at transmission and the number of transmitted lineages, in line with theoretical predictions [48]. However, the ability to select best-fitting phylogenies when exact matches were not possible highlights both the model’s robustness and its limitations, particularly given the challenges of capturing the full complexity of host-pathogen interactions during transmission and additional uncertainties in individual data accuracy.

Our study has several limitations: (1) the model did not incorporate natural selection, which in [7] was assumed to act by preferentially transmitting variants more similar to those that initiated the infection, thereby reducing the diversity of variants available for onward transmission. However, if natural selection does occur, any reduction in diversity within the transmitter’s virus inoculum would be offset by an increased per-virion acquisition probability, allowing more virions and variants to be transmitted overall [7]. (2) Significant gaps exist in understanding how genetic diversity differs and relates between different physiological compartments over time. This could potentially affect our findings, as most genetic data comes from blood samples rather than sexual bodily fluids, which may not fully capture the nature of viral populations involved in sexual transmission events. (3) We observed correlations among some fitted parameters, suggesting partial non-identifiability; however, disentangling their individual contributions would require additional experimental data. (4) Our phylodynamic approach assumes that each founder variant maintains a recognisable lineage from transmission to sampling, not accounting for recombination. But, in reality, recombination can obscure the true number of founder variants [49].

In conclusion, we present a mechanistic model of HIV transmission that reconciles key epidemiological observations through three biologically grounded mechanisms: intermittent susceptibility, stage-dependent infectivity, and target cell limitation. The model provides insight into the factors underlying differences in transmission risk between populations, and its ability to recapitulate independent observations—including the very low transmission risk from individuals with suppressed viral loads—supports its utility as a principled framework for understanding HIV transmission biology and evaluating prevention strategies.

## Methods

### Transmitter characterization

Infectious untreated individuals progress through three stages of infection: early, asymptomatic, and late, lasting *τ_E_*, *τ_A_*(*v_A_*), and *τ_L_* respectively. A randomly selected transmitter is characterized by their viral load in the genital mucosa in each stage, V = (*V_E_, V_A_, V_L_*), taking independent values v = (*v_E_, v_A_, v_L_*), which occur with probability:

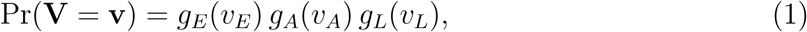

where *g_E_*, *g_A_* and *g_L_* are the relative frequencies of viral loads in the transmitting population at each stage, respectively. The value of *v_A_* fixes the duration of the asymptomatic stage *τ_A_*(*v_A_*) [8], and we assume that *τ_E_*and *τ_L_* are fixed (i.e., independent of the viral load). We assume that each exposure occurs at a random time Θ since the transmitter became infected, uniformly distributed throughout the infectious period:

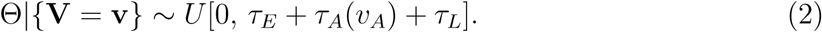

The stage of the transmitter’s infection, *S* = *s*(V, Θ) ∈ {*E, A, L*}, is defined by:

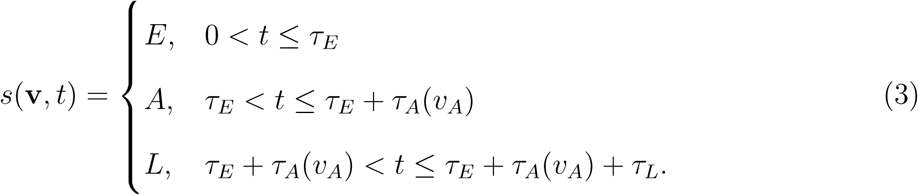

Since Θ is uniform, the probability that the transmitter is in stage *s* ∈ {*E, A, L*} is:

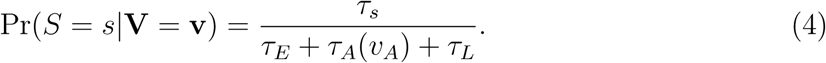

### Intermittent susceptibility model

In the intermittent susceptibility model, HIV infection can only occur during a small fraction of sexual exposures—specifically those that occur when conditions are permissive for transmission. Our intermittent susceptibility model follows the same structure as described elsewhere [7], with two modifications: first, viral loads vary between individuals in the population in all infection stages, consistent with epidemiological data [50, 51]. Second, for mathematical tractability, we approximate the number of virions transmitted to the exposed partner using a Poisson distribution (rather than a binomial distribution) because the viral load in the genital mucosa, *v*, is large and the per-virion acquisition probability, *p*, is small. In this model, the probability that *N*_1_ virions establish infection *during a single sexual exposure*, assuming conditions are permissive for infection, is given by:

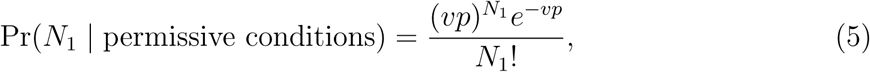

for *N*_1_ = 0, 1, 2, 3 … , *v*. Accounting for the additional possibility that conditions are not permissive, the probability that infection does not occur is:

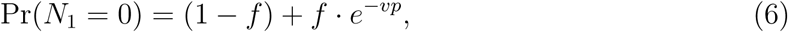

where *f* is the probability of permissive conditions for infection. Similarly, the probability that *Y* genetic variants are transmitted during a single sexual exposure resulting in infection is given by:

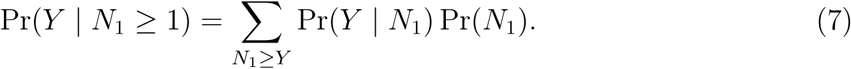

We calculate the probability that *N*_1_ virions establish infection *for any exposure* under permissive conditions as:

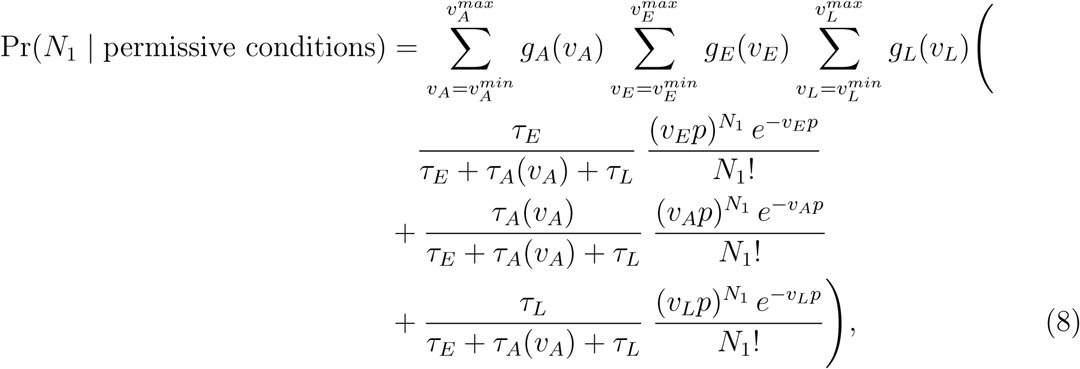

where *N*_1_ ≥ 0. The expected probability of no transmission *for any exposure* is therefore:

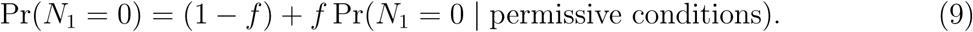

The probability of *Y* variants being transmitted during a single transmission event is then:

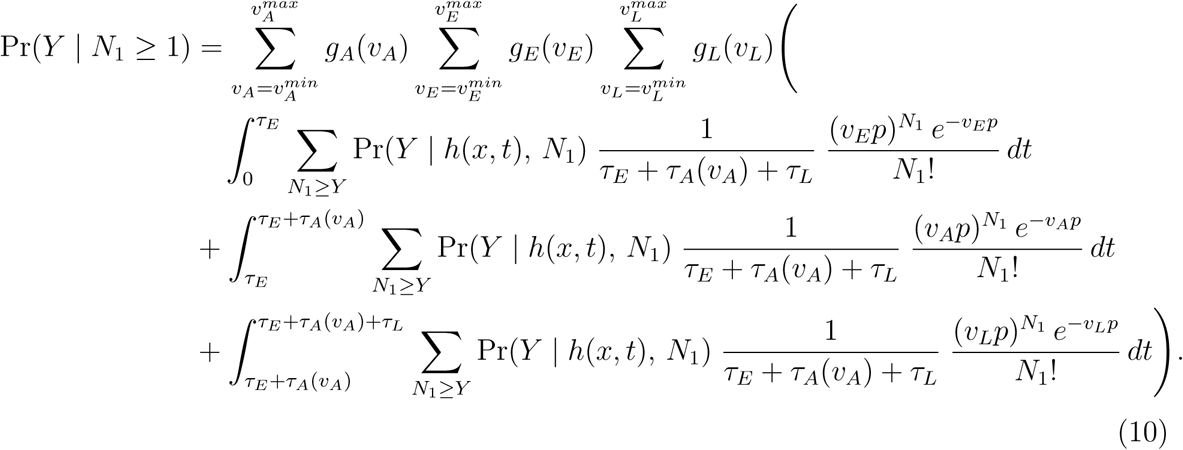

In the equation above, we track the distribution of the number of genetic variants avail­able for transmission as *h*(*x, t*). This expression represents the proportion of the *x^th^* most abundant genetic variant at time *t* years after the transmitting partner became infected and is characterised using the following approximation [7]:

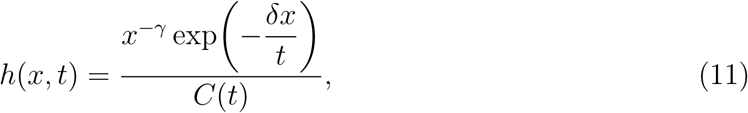

for *x* = 1, 2, 3*, … , x_max_*, where *x_max_* is the maximum number of distinct genetic variants observed in any individual at any single time, *γ* = 0.583 and *δ* = 0.563 are derived from the same data, and *C*(*t*) is a normalising constant ensuring 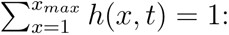

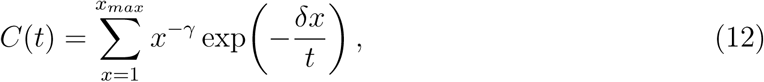

previously parameterised by [7] using data from [52].

### Intermittent susceptibility and stage-dependent virus infectivity model

We extend our updated intermittent susceptibility model by relaxing the assumption that the transmitter’s per-virion infectivity remains constant throughout their infectious period. To this end, we introduce stage-dependent per-virion transmission probabilities. The expressions are all identical to above, with *p* replaced by the stage-specific value *p_s_*, where *s* ∈ {*E, A, L*} for early, asymptomatic, and late stages of the transmitter, respectively.

### Intermittent susceptibility, stage-dependent virus infectivity and target-cell-limited model

Next, we extend our model by assuming that a finite number of target cells is initially available for infection at the exposure site. The number of virions transmitted to the exposed partner remains Poisson-distributed after exposure under permissive conditions for infection:

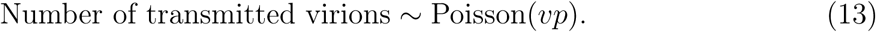

Each transmitted virion independently attaches to one of the *c* target cells in the genital mucosa, so the number of transmitted virions per cell is:

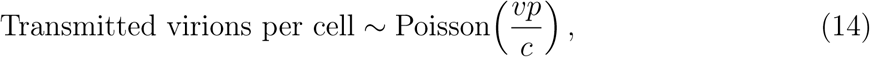

with the probability that each cell becomes infected as:

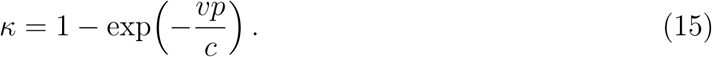

Assuming that the number of target cells available for infection is a random variable that follows a Poisson distribution with parameter *c*, then the number of infected cells after exposure is:

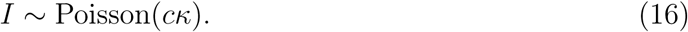

In this model, stage-dependent differences in virus infectivity are captured by the cell establishment probability *r_s_* rather than the per-virion transmission probability *p_s_*, which remains constant across stages (*p*). This reflects the assumption that stage-dependent variation in infectivity arises at the level of cellular establishment of systemic infection rather than at the level of initial virion transmission. Thus, we assume that *N*_2_, the number of infected cells that establish infection in the exposed partner, is binomially distributed with a stage-dependent probability *r_s_*, where *s* ∈ {*E, A, L*} for early, asymptomatic, and late stages of the transmitter, respectively:

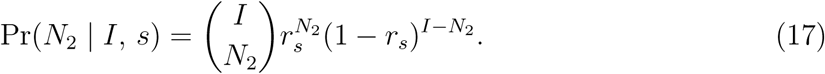

The probability that *N*_2_ cells (via exactly *N*_2_ virions) establish infection in a single exposure during a transmitting partner’s infection stage is then given by:

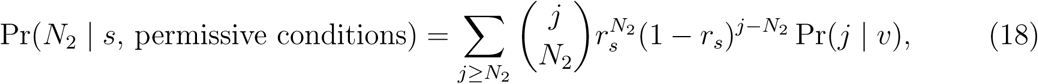

for *N*_2_ ≥ 0, where:

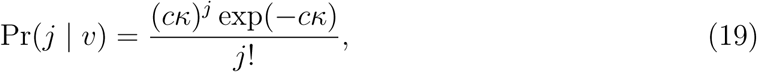

is the probability that *j* cells are initially infected in the exposed partner, given a viral load. The average probability of no transmission is:

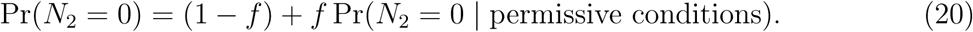

The expected probability that *N*_2_ cells (via exactly *N*_2_ virions) establish infection for any exposure under permissive conditions is:

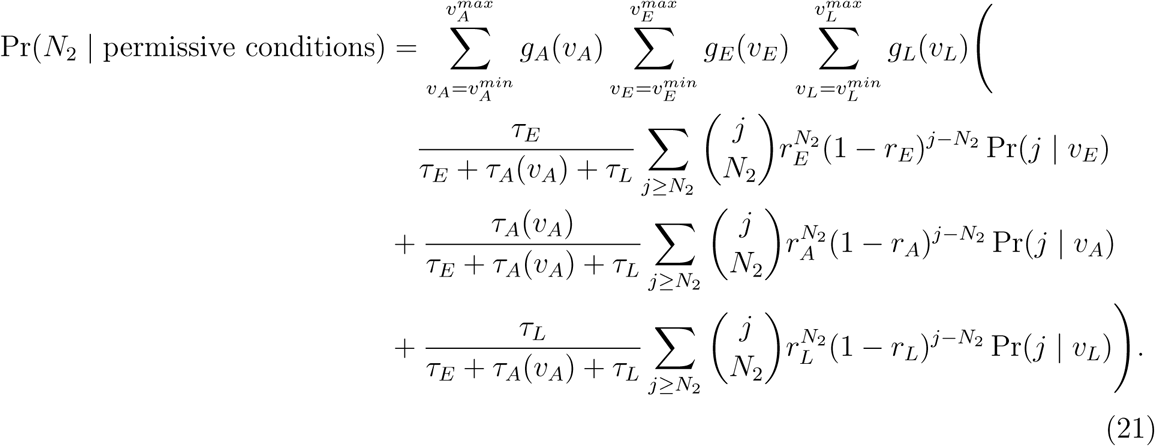

The joint probability of *N*_2_ cells (via exactly *N*_2_ virions) and *Y* variants establishing infection is given by:

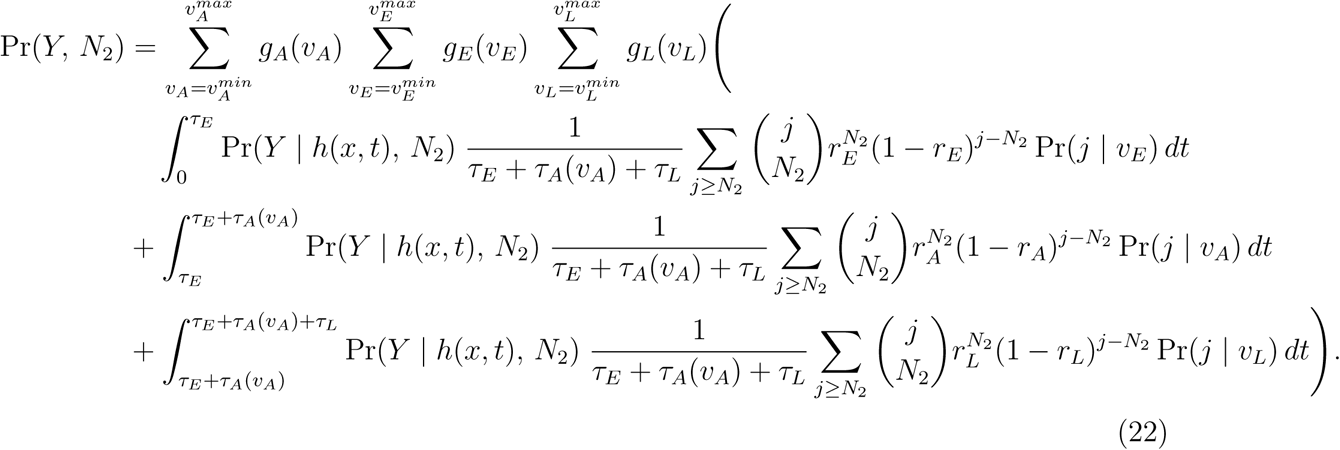

For this model, we can also calculate the probability of transient infection during stage *s*—That is, exposures where cell infection occurs but the viral population is extinguished stochastically and before IgG seroconversion:

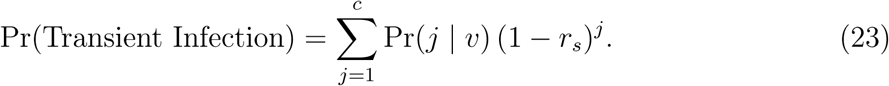

### Model suite

We analysed seven models representing different combinations of the mechanisms described above (**Supplementary Table 4**):

- M1: Intermittent susceptibility model (as above).
- M2: Stage-dependent virus infectivity model (M3 with *f* = 1).
- M3: Intermittent susceptibility and stage-dependent virus infectivity model (as above).
- M4: Target-cell-limited model (M7 with *f* = 1 and *r_s_* = *r*).
- M5: Intermittent susceptibility, target-cell-limited model (M7 with *r_s_* = *r*).
- M6: Stage-dependent virus infectivity and target-cell-limited model (M7 with *f* = 1).
- M7: Intermittent susceptibility, stage-dependent virus infectivity and target-cell-limited model (as above).

Full mathematical derivations of the models, including a unified Poisson framework, are provided in the **Supplementary Material**.

### Parameterisation and model fitting

#### Viral loads

We derived the viral load frequency during the early stage of infection, *g_E_*(*v_E_*), by fitting a normal distribution to averages of viral load quantiles across the first five Fiebig stages, with each stage contributing in proportion to its duration [51].

We assumed an individual’s viral load during the asymptomatic stage remains at a set point, with the viral load weights across the transmitting population, *g_A_*(*v_A_*), following a skew-normal distribution [7].

We derived the viral load weights in the late stage of infection, *g_L_*(*v_L_*), by fitting a log-normal distribution to viral load counts collected before AIDS onset from a cohort of men who have sex with men [50], with a median viral load of 103,900 copies per mL. The same distribution was used for both MSM and heterosexual transmission (as the median viral loads for late-stage index partners from HIV-discordant heterosexual couples was similar at 112,600 copies per mL) [53].

#### Infection stage durations

We estimated the duration of the early stage, *τ_E_*, using the distribution from [13] (mean 52 days, 95% CI: 17–207) as a prior (**Supplementary Table 4**). The duration (in years) of an individual transmitter’s asymptomatic stage is a function of their set-point viral load, *v_A_*:

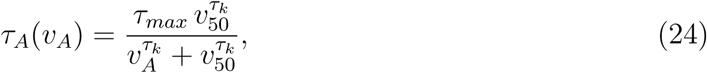

where *τ_max_* is the maximum duration (25.4 years), *v*_50_ is the viral load at which the duration is half its maximum (3,058 copies per mL), and *τ_k_* is the steepness of the decrease in duration as a function of viral load (0.41) [8]. This parameterisation accounts for the fact that individuals with higher set-point viral loads progress more quickly through the asymptomatic stage and are therefore underrepresented in cross-sectional samples of asymptomatic individuals at any given time. We therefore rescale:

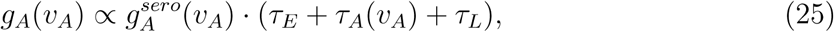

following [8] where 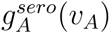 is the frequency of viral loads in a cohort of seroconverters . The duration of the late stage, *τ_L_*, was assumed to be constant across all transmitters and fixed at ten months [13].

### Number of sexual exposures per year

We estimated annual sexual exposures between uninfected individuals and their long-term asymptomatically infected sexual partners, using a normal distribution with a mean of 107 exposures and standard deviation of 23.5 as a prior, based on reported intercourse frequency among monogamous, heterosexual, HIV-1-discordant couples from Rakai, Uganda [54]. We applied the same prior to our MSM analysis, as the estimated annual mean intercourse frequency among men who have sex with a main sexual male partner was similar at 102 exposures [55].

### Additional parameters

We assumed uniform priors for all remaining model probability parameters: *p* (probability of per-virion acquisition, either stage-dependent or constant), *f* (probability of permissive conditions for infection), and *r* (probability that an infected cell establishes systemic in­fection, either stage-dependent or constant) on the interval [0, 1]. Parameter *c* (number of target cells available for initial infection in the exposed partner) was assigned a prior on the interval [1, 1000].

### Model fitting

We fit each model independently via Markov chain Monte Carlo to six observations for heterosexual HIV transmission: (1) the per sexual exposure probability of HIV acquisition during the asymptomatic stage for heterosexual transmission [4], (2) the relative hazard of HIV acquisition during the early versus asymptomatic stage [13] estimated in a cohort of heterosexual partners, (3) the relative hazard of HIV acquisition during the late versus asymptomatic stage for heterosexual partners [13], (4) the per sexual exposure probability of multiple variant transmission during heterosexual transmission [6], (5) the relative hazard of HIV multiple founder variants during the early versus asymptomatic stage [56], and (6) the annual HIV transmission rate stratified by set-point viral load between serodiscordant heterosexual partners [12] (**Supplementary Table 5**). We removed observation five from the fitting procedure in a sensitivity analysis.

For observation six, we defined the annual individual risk after *ε* exposures with an individual with an asymptomatic viral load, *v_A_*, by:

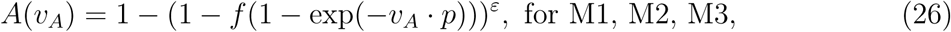

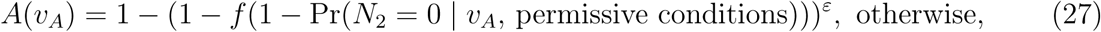

where,

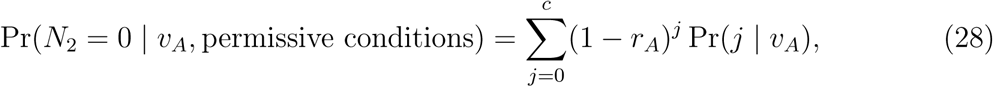

and Pr(*j* | *v_A_*) is the Poisson distribution of initially infected cells as defined earlier in Equation 19.

Model fitting was performed using Markov chain Monte Carlo. We ran four independent chains, each with 50,000 iterations. We discarded the first 25% of iterations as burn-in to exclude samples drawn before convergence, and thinned the chains by retaining every 75th iteration to reduce autocorrelation. The combination of the four chains yielded 2,000 posterior samples. We assessed convergence by ensuring that the multivariate potential scale reduction factor (Gelman-Rubin convergence diagnostic) was smaller than 1.1 and that the effective sample size for each parameter was higher than 200.

After fitting each model to the data from heterosexual transmission, we refit each model for men who have sex with men (MSM) transmission. Specifically, we made the following updates to the six observations: (1) a per-exposure per-stage acquisition probability that was 4.16 times higher in MSM than in heterosexuals, using the average of the ratios of anal intercourse acquisition probabilities [4], (2) the relative hazard of HIV acquisition during the early versus asymptomatic stage and (3) during the late versus asymptomatic stage remained the same as the heterosexual data [13], (4) a higher per sexual exposure probability of multiple variant transmission observed during MSM transmission [6], (5) the relative hazard of HIV multiple founder variants during the early versus asymptomatic stage remained the same [56], (6) the annual HIV transmission rate was 1.57 times higher in MSM than in heterosexuals. This value corresponds to the relative hazard of transmission rates per year by exposure group, calculated from the reported US CDC national HIV incidence and prevalence rates in the United States for 2015 (**Supplementary Table 5**).

### Individual-level phylodynamic analysis

We combined our best-fitting transmission model with a coalescent-based phylodynamic model to estimate the time since infection and viral load at transmission for 48 individual transmission pairs. For each pair, we drew values from the uncertainty ranges of these transmission characteristics as inputs to our transmission model, which predicted the number of founding virions and variants. These predictions, together with sampling times and sequence data, were used to simulate transmission trees under a coalescent framework, from which we inferred phylogenies and computed five summary statistics (see **Supplementary Methods** for details of the individual-level analysis, including summary statistic definitions). We used Approximate Bayesian Computation Sequential Monte Carlo (ABC-SMC) to fit the phylodynamic model to each transmission pair, estimating posterior distributions by comparing simulated and observed phylogenetic trees.

## Data Availability

All data produced are available online at: https://github.com/Chjulian/HIV_reconciliation

## Code Availability

Code for running the model and generating the distribution of variants in transmitters is accessible at: https://github.com/Chjulian/HIV_reconciliation.

## Acknowledgements

This project has received funding from the European Research Council (ERC) under the European Union’s Horizon 2020 research and innovation programme (grant agreement 757688). Katrina Lythgoe was supported by the Wellcome Trust and the Royal Society (107652/Z/15/A), Wellcome Trust (227438/Z/23/Z), MRC (MR/V035304/1), and the Li Ka Shing Foundation.

