## Supplementary Text for "Barriers in recipient partners cause frequent transient HIV infections and explain transmission risk under viral sup­pression"

### Supplementary material for: Accounting for barriers to HIV infection in the recipient partner reveals frequent transient infections and explains transmission risk under viral suppression

Katherine E. Atkins[1,2,3], Tibor Antal[4], Katrina Lythgoe[5], Roland Regoes[6], Robin N. Thompson[7], Stéphane Hué[2,3], and Ch. Julián Villabona-Arenas[3,4]

[1]Usher Institute, University of Edinburgh, Edinburgh, United Kingdom
[2]Centre for Mathematical Modelling of Infectious Diseases, London School of Hygiene and Tropical Medicine, London, United Kingdom
[3]Department of Infectious Disease Epidemiology & Dynamics, London School of Hygiene and Tropical Medicine, London, United Kingdom
[4]School of Mathematics and the Maxwell Institute for Mathematical Sciences, University of Edinburgh, Edinburgh, United Kingdom
[5]Big Data Institute, University of Oxford, United Kingdom
[6]Integrative Biology, ETH Zurich, 8092 Zurich, Switzerland
[7]Mathematical Institute, University of Oxford, Oxford, United Kingdom

**2 Supplementary Tables** **17**

**3 Supplementary Figures** **22**

### 1 Supplementary methods

#### 1.1 Derivation of the models

##### 1.1.1 Notation

| | |
|---|---|
| Viral load | $\mathbf{V} = (V_E, V_A, V_L), \mathbf{v} = (v_E, v_A, v_L)$ |
| Time since infection | $\Theta, t$ |
| Number of established virions | $N_1, n$ |
| Number of established cells | $N_2, n$ |
| Mean number of established infections | $\lambda_1, \lambda_2$ |
| Stage of transmitter | $S, s \in \{E, A, L\}$ |
| Number of distinct genetic variants | $Y, y$ |
| Number of infected cells | $I$ |

##### 1.1.2 Number of established infections

**Intermittent susceptibility model.** Given $v \gg 1$ virions, the number of established infections (for permissive conditions for infection) is approximately $N_1 \sim \text{Poisson}(pv)$, since each virion becomes established independently with probability $p \ll 1$. Since $v \in \{v_E, v_A, v_L\}$ according to the stage $S$ the transmitter is in, we get:

$$\Pr(N_1 = n | S = s, \mathbf{V} = \mathbf{v}) = \frac{(pv_s)^n}{n!} e^{-pv_s}. \tag{1}$$

**Target-cell-limited model.** Again, we have $v \gg 1$ transmitted virions, and the number of established infections (for permissive conditions) is approximately $N_1 \sim \text{Poisson}(pv)$, since each virion becomes established independently with probability $p \ll 1$. These $N$ transmitted virions then independently and uniformly infect the fixed $C = c$ cells in the genital mucosa. Hence we can split the virions into the corresponding cells they infect, so each cell receives $\text{Poisson}(pv/c)$ virions independently. This means that each cell is independently infected with at least one virion with probability:

$$\kappa = 1 - e^{-pv/c}, \tag{2}$$

and the total number of infected cells is:

$$I|\{C = c\} \sim \text{Binomial}(c, \kappa). \tag{3}$$

Now assume that the number of cells in the genital mucosa is not fixed but $C \sim \text{Poisson}(c)$, in which case the number of infected cells is:

$$I \sim \text{Poisson}(c\kappa). \tag{4}$$

We can obtain this result from conditioning and using the previous binomial result as:

$$\begin{aligned}
\Pr(I = a) &= \sum_{b \geq a} \Pr(I = a|C = b) \Pr(C = b) \\
&= \sum_{b \geq a} \binom{b}{a} \kappa^a (1 - \kappa)^{b-a} \frac{c^b}{b!} e^{-c} \\
&= \frac{e^{-c}}{a!} (c\kappa)^a \sum_{b \geq a} \frac{(c(1 - \kappa))^{b-a}}{(b - a)!} \\
&= \frac{(c\kappa)^a}{a!} e^{-c\kappa},
\end{aligned} \tag{5}$$

that is $I \sim \text{Poisson}(c\kappa)$.

Not all of these $I$ cells establish infections in the exposed partner, but each independently does that with probability $r$, which will be stage dependent. Hence the number of established cells $N_2 \sim \text{Poisson}(c\kappa r)$ (with permissive condition) due to the splitting property, so for example $\Pr(N_2 = 0) = e^{-c\kappa r}$.

Let us specify now for each stage $s \in \{E, A, L\}$, that we have stage dependent $v_s, \kappa_s = 1 - e^{-pv_s/c}, r_s$. Hence conditioning for stage $S$ and viral loads $\mathbf{V}$ the mass function of the number of established infections, $N_2$, becomes:

$$\Pr(N_2 = n|S = s, \mathbf{V} = \mathbf{v}) = \frac{(c\kappa_s r_s)^n}{n!} e^{-c\kappa_s r_s}. \tag{6}$$

**Poisson established infections.** Hence, both of the above models predict a Poisson number of established infections:

$$N_i|\{S = s, \mathbf{V} = \mathbf{v}\} \sim \text{Poisson}(\lambda_i), \tag{7}$$

for $i \in \{1, 2\}$; they only differ in their parameters:

$$\lambda_1 = pv_s, \quad \lambda_2 = cr_s(1 - e^{-pv_s/c}). \tag{8}$$

Since stage $S$ is determined by $\Theta$ and $\mathbf{V}$, we can equivalently condition on $\Theta$ here:

$$N_i|\{\Theta = t, \mathbf{V} = \mathbf{v}\} \sim N_i|\{S = s(\mathbf{v}, t), \mathbf{V} = \mathbf{v}\} \sim \text{Poisson}(\lambda_i), \tag{9}$$

with:

$$\lambda_1 = pv_{s(\mathbf{v},t)}, \quad \lambda_2 = cr_{s(\mathbf{v},t)}(1 - e^{-pv_{s(\mathbf{v},t)}/c}), \tag{10}$$

and the function $s(\mathbf{v}, t)$ is as defined in the main text.

##### 1.1.3 Unconditional distribution of established infections

Hence conditioned on $v_E, v_A, v_L$, the number of established infections is:

$$\begin{aligned} \Pr(N_i = n|\mathbf{V} = \mathbf{v}) &= \sum_{s \in \{E,A,L\}} \Pr(N_i = n|S = s, \mathbf{V} = \mathbf{v}) \Pr(S = s|\mathbf{V} = \mathbf{v}) \\ &= \sum_{s \in \{E,A,L\}} \frac{\tau_s}{\tau_E + \tau_A(v_A) + \tau_L} \frac{\lambda_i^n}{n!} e^{-\lambda_i}, \end{aligned} \tag{11}$$

where we used the stage probability from the main text and (1). Taking the expectation over the number of virions we get the mass function of the number of established infections:

$$\begin{aligned} \Pr(N_i = n) &= \sum_{v_E, v_A, v_L} \Pr(N_i = n|\mathbf{V} = \mathbf{v}) \Pr(\mathbf{V} = \mathbf{v}) \\ &= \sum_{v_E, v_A, v_L} g(\mathbf{v}) \sum_{s \in \{E,A,L\}} \frac{\tau_s}{\tau_E + \tau_A(v_A) + \tau_L} \frac{\lambda_i^n}{n!} e^{-\lambda_i}, \end{aligned} \tag{12}$$

which appears expanded in the main text.

##### 1.1.4 Number of genetic variants

The main quantity of interest is the number $Y$ of distinct genetic variants transmitted by a transmitter. Let us condition again on the viral loads in each stage, $v_E, v_A, v_L$, and on the time since infection $\Theta = t$ of the transmitter. The number of established virions $N$ in both models are Poisson distributed as in (9), with parameter $\lambda$. More precisely, $N \in \{N_1, N_2\}$ with corresponding $\lambda \in \{\lambda_1, \lambda_2\}$, but we drop the indices now for brevity.

Each established virion is independently of variant $x$ with a time since infection $t$ dependent probability:

$$h(x,t) = \frac{1}{C(t)} x^{-\gamma} e^{-\delta x/t}, \tag{13}$$

with $\gamma = 0.583, \delta = 0.563$, and:

$$C(t) = \sum_x x^{-\gamma} e^{-\delta x/t}, \tag{14}$$

over the range of $x$, so $\sum_x h(x,t) = 1$. If $x \geq 1$ but unbounded (in practice a finite range is fine due to the exponential decay), then $C(t) = \mathrm{Li}_\gamma(e^{-\delta/t})$ can be expressed in terms of the polylogarithm function.

Hence the number of established virions of variant $x$ is also a Poisson:

$$I_x \sim \mathrm{Poisson}(\lambda h(x,t)), \tag{15}$$

due to the splitting property. The number of variants transmitted is:

$$Y = \sum_x {}_{I_x > 0}, \tag{16}$$

and its conditional mass function is:

$$\Pi_{t,v_s}(y) = \Pr(Y = y | \mathbf{V} = \mathbf{v}, \Theta = t) = \Pr\left( \sum_x {}_{I_x > 0} = y | \mathbf{V} = \mathbf{v}, \Theta = t \right), \tag{17}$$

where $s = s(\mathbf{v}, t)$. We use index $v_s$ for $\Pi$ since only that viral load enters via $\lambda$. Hence for a given $\mathbf{v}, t$ one just needs to generate one Poisson random variable for each $x$ (up to some bound) to calculate $Y$. With the above method, one spares assigning each virion to a variant, as that is included in the Poisson already.

Let us abbreviate the conditional probability as $\Pr^*(\cdot) = \Pr(\cdot|\mathbf{V} = \mathbf{v}, \Theta = t)$ for now. For $y = 0$ we get:

$$\begin{aligned}\Pi_{t,v_s}(0) &= \overset{*}{\Pr}(I_x = 0,\ \forall x) = \prod_x \overset{*}{\Pr}(I_x = 0) \\ &= \prod_x e^{-\lambda h(x,t)} = e^{-\sum_x \lambda h(x,t)} = e^{-\lambda}, \end{aligned} \tag{18}$$

where we used the independence of $I_x$ of different indices. This is expected as we have no variants if and only if there are no established virions, which happens with probability $e^{-\lambda}$.

Similarly:

$$\begin{aligned}\Pi_{t,v_s}(1) &= \sum_x \overset{*}{\Pr}(I_x > 0, I_y = 0\ \forall y \neq x) = \sum_x e^{-\lambda} \frac{\Pr^*(I_x \geq 1)}{\Pr^*(I_x = 0)} \\ &= e^{-\lambda} \sum_x \frac{1 - e^{-\lambda h(x,t)}}{e^{-\lambda h(x,t)}} = e^{-\lambda} \sum_x (e^{\lambda h(x,t)} - 1), \end{aligned} \tag{19}$$

and:

$$\begin{aligned}\Pi_{t,v_s}(2) &= \sum_{x,y,x<y} \overset{*}{\Pr}(I_x, I_y > 0, I_z = 0\ \forall z \neq x, y) \\ &= \sum_{x,y,x<y} e^{-\lambda} \frac{\Pr^*(I_x \geq 1)}{\Pr^*(I_x = 0)} \frac{\Pr^*(I_y \geq 1)}{\Pr^*(I_y = 0)} \\ &= e^{-\lambda} \sum_{x,y,x<y} (e^{\lambda h(x,t)} - 1)(e^{\lambda h(y,t)} - 1). \end{aligned} \tag{20}$$

Analogously:

$$\Pi_{t,v_s}(y) = e^{-\lambda} \sum_{x_1 < \cdots < x_y} (e^{\lambda h(x_1,t)} - 1) \ldots (e^{\lambda h(x_y,t)} - 1). \tag{21}$$

To get rid of the conditioning, we take expectations, first over time and then over viral load, $\Pr(Y=y) = \{[\Pr(Y=y|\Theta)|\mathbf{V}]\}$, to obtain the distribution of the number of distinct genetic variants:

$$\Pr(Y=y) = \sum_{v_E, v_A, v_L} \frac{\int \Pi_{t,v_s}(y)dt}{\tau_E + \tau_A(v_A) + \tau_L} \times g(\mathbf{v}), \tag{22}$$

where the integration is over $[0, \tau_E + \tau_A(v_A) + \tau_L]$. Here, $\Pi_{t,v_s}(y)$ with $s = s(\mathbf{v}, t)$ is given by (17), $I_x \sim \text{Poisson}(\lambda h(x,t))$, $h(x,t)$ is given by (13), and finally $\lambda \in \{\lambda_1, \lambda_2\}$ depending on the model and given by (10).

Note that we can write out the above integral as:

$$\int_0^{\tau_E} \Pi_{t,v_E}(y)dt + \int_{\tau_E}^{\tau_E+\tau_A(v_A)} \Pi_{t,v_A}(y)dt + \int_{\tau_E+\tau_A(v_A)}^{\tau_E+\tau_A(v_A)+\tau_L} \Pi_{t,v_L}(y)dt, \tag{23}$$

which is how it is written in the main text. Let us call these above integrals $i_E, i_A, i_L$. The first integral $i_E$ depends only on $v_E$ (fixed limits $[0, \tau_E]$), and $i_A$ depends only on $v_A$. However, $i_L$ depends on both $v_L$ (through $\lambda$ in $\Pi$) and $v_A$ (through the integration limits $[\tau_E + \tau_A(v_A),\ \tau_E + \tau_A(v_A) + \tau_L]$, since $h(x,t)$ depends on absolute time $t$). Therefore $i_L = i_L(v_A, v_L)$, and we can rewrite the distribution of the number of distinct genetic variants as:

$$\Pr(Y=y) = \Theta \sum_{v_E} g(v_E) i_E + \sum_{v_A} g(v_A) \frac{i_A}{\tau_E + \tau_A(v_A) + \tau_L} + \Theta \sum_{v_L} g(v_L)\, i_L, \tag{24}$$

where:

$$\Theta = \frac{1}{\tau_E + \tau_A(V_A) + \tau_L} = \sum_{v_A} \frac{g(v_A)}{\tau_E + \tau_A(v_A) + \tau_L}, \tag{25}$$

is the mean inverse duration time of the infection.

**Mean number of variants.** The mean of the number of established genetic variants is somewhat simpler:

$$(Y|\mathbf{V} = \mathbf{v}, \Theta = t) = \sum_x \Pr(I_x > 0) = \sum_x (1 - e^{-\lambda h(x,t)}), \tag{26}$$

and by taking its expectation over $\Theta$ then over $\mathbf{V}$ we obtain:

$$(Y) = \sum_{v_E, v_A, v_L} \frac{\sum_x \int (1 - e^{-\lambda h(x,t)}) dt}{\tau_E + \tau_A(v_A) + \tau_L} \times g(\mathbf{v}), \tag{27}$$

where the integration is over $[0, \tau_E + \tau_A(v_A) + \tau_L]$.

#### 1.2 Individual-level phylodynamic analysis

##### 1.2.1 Data

We collated 48 transmission pairs of HIV-infected individuals with documented linkage, which included viral load counts from the transmitter's partner at sampling, HIV sequences from both individuals, and transmission timelines. Transmission timelines included the time since infection of the transmitter partner (quantitative or qualitative) and the relative time from transmission to sampling from both individuals. We classified transmitter individuals as early transmitters if most of their infection duration at transmission fell within a 90-day window (n=8), or as asymptomatic transmitters otherwise (n=40). We did not classify individuals as late-stage (pre-AIDS) transmitters, as this typically requires additional CD4 count data.

We used approximate Bayesian computation based on Sequential Monte Carlo sampling (ABC-SMC) to estimate parameters for which direct measurements were unavailable. Of the eight early transmitters, four individuals had an uncertain time since infection at transmission. Of the 40 asymptomatic transmitters, 39 individuals had an uncertain time since infection at transmission. For asymptomatic transmitters with unknown or right-censored time since infection at transmission, we used bounds based on the minimum documented time (90 days if unknown) and the maximum documented time across the data (approximately 11 years).

Early transmitters had a median delay from transmission to sampling of 46 days (maximum 128 days). Given that viral load is highly dynamic during this stage [1], we estimated viral load at transmission for all eight early transmitters, assuming it was bounded within $\pm 1.5$ $\log_{10}$ copies/mL of the observed value at sampling. Asymptomatic transmitters had a median delay from transmission to sampling of 66 days (maximum 299 days). Given that set-point viral load (SPVL) typically remains stable during this stage, we assumed viral load at transmission equalled the measured SPVL [2]. In summary, we applied ABC-SMC to estimate posterior distributions of time since infection at transmission for 43 transmitter partners (four early, 39 asymptomatic) and of viral load at transmission for all eight early transmitters.

##### 1.2.2 Prior distributions

For early transmitters, the prior distribution of the viral load at transmission, $V$, is given by fitting a normal distribution to averages of viral load quantiles across the first five Fiebig stages, with each stage contributing in proportion to its duration [1]. The prior distribution of time since infection at transmission, $t$, is based on the estimated duration of the acute phase by [3].

For asymptomatic transmitters, $V$ corresponds to SPVL, and we calculated the joint prior distribution ($\pi$) of $V$, $t$, and, $\Delta_A$ (relative time to sampling of the transmitter partner) for an individual $i$, as follows:

$$\pi_i(\theta) = P(V, t, \Delta_A) = \alpha[\text{transmission probability} \times \text{survival probability}]. \tag{28}$$

This prior probability tells us the probability that an HIV-infected individual with a given SPVL lived long enough to reach time $t$, transmitted the virus, and survived until the sampling time. We modelled the probability of transmission per person per year using a Poisson process, where the transmission rate, $\beta$, was a function of set-point viral load (SPVL) [2]:

$$1 - \exp(-\beta(V)t), \tag{29}$$

where $\beta(V)$ is defined as:

$$\beta(V) = \frac{\beta_{max} V^{\beta_k}}{(V^{\beta_k} + \beta_{50}^{\beta_k})}, \tag{30}$$

with parameters $\beta_{max} = 0.317$ per year, $\beta_{50} = 13{,}938$ copies per mL of peripheral blood and $\beta_k = 1.02$. We defined $Z$ as the time (in years) from initial infection to death from untreated

HIV. Given that the infectious period comprised three stages, Z was calculated as the sum of their durations:

$$Z = Z_{early} + Z_{Asymptomatic} + Z_{Late}. \tag{31}$$

We assumed that:

$$Z_{\text{early}} \sim \text{log-normal}(\mu_{\text{log}} = -1.95,\ \text{sd}_{\text{log}} = 0.69) \qquad [3], \tag{32}$$

$$Z_{\text{Asymptomatic}} \sim \Gamma(\mu = D(V),\ \text{shape} = 3.46) \qquad [2], \tag{33}$$

where:

$$D(V) = \frac{D_{max} \cdot D_{50}^{D_k}}{V^{D_k} + D_{50}^{D_k}}, \tag{34}$$

with $D_{max} = 25.4$ years, $D_{50} = 3{,}058$ copies/mL, and $D_k = 0.41$, and:

$$Z_{Late} = 10/12 \quad [3]. \tag{35}$$

Given $\Delta_A$, the delay between transmission and sampling of the transmitter, we calculated the empirical cumulative distribution for $Z$, using random draws from the three underlying distributions and defined the survivorship function:

$$S(V, E) = 1 - F(V, t, \Delta), \tag{36}$$

where:

$$E = \max(t, t + \Delta). \tag{37}$$

Thus, the joint prior distribution of $V$, $t$, and $\Delta_A$ for an individual $i$, is given by:

$$\pi_i(\theta) = P(V, t, \Delta_A) = \alpha[(1 - \exp(-\beta(V)t))\ S(V, E)]. \tag{38}$$

##### 1.2.3 Sequence data simulation

For each transmission pair, we simulated time-calibrated transmission trees using VirusTreeSimulator (`github.com/PangeaHIV/VirusTreeSimulator`), employing a within-host effective population size as described in [4]. Transmission trees were parameterised by the transmitter's time since infection at transmission ($t$), the number of transmitted virions and variants in each partner, sampling times, and the sampling times and number of unique sequences (number of simulated tips) from each partner.

For the transmitter partner, we drew the number of transmitted virions ($N_B$) and variants ($Y_B$) from a population-level joint probability distribution (assuming approximately 20% of transmission events involve multiple founder variants). For the exposed partner, we drew the number of founding virions ($N_A$) and genetic variants ($Y_A$) from the joint probability distribution derived from our best-fitting transmission model. Given the observed correlations among parameters (**Supplementary Figure 2**) and to reduce computational burden, we fixed $c = 16$ and set other parameters to their posterior estimates conditional on this value.

Simulated transmission trees were converted to branch lengths in units of substitutions per site by assuming a strict molecular clock with a substitution rate matching that of the empirical genomic region. We simulated nucleotide sequence evolution along these trees using AliSim [5, 6] in IQ-TREE [7], with sequence lengths matching the empirical alignments.

###### 1.2.4 Phylogenetic inference

For each transmission pair, we aligned empirical HIV sequences using MAFFT [8] and inferred maximum-likelihood trees using 100 independent searches in IQ-TREE [7], with ModelFinder [9] for substitution model selection. Empirical trees were rooted using outgroup sequences identified via BLAST and the Los Alamos HIV database, then excluded from downstream analyses as in [10]. For simulated HIV sequences, we inferred maximum-likelihood trees using the same substitution model as the empirical data.

###### 1.2.5 Model fitting via Approximate Bayesian Computation

We inferred a phylogeny ($y$) for each set of simulated HIV sequences. This allowed us to compare simulated and observed data ($d$) via summary statistics computed from the phylogenies.

Model fitting was performed using Approximate Bayesian Computation Sequential Monte Carlo (ABC-SMC). We used the algorithm proposed by [11], as in [12], with two key modifications. First, we introduced separate tolerance levels $\varepsilon^1, \ldots, \varepsilon^X$ for each summary statistic. Second, instead of a distance metric $\rho(\cdot, \cdot)$ that combines all summary statistic comparisons into a single scalar value, we adopted a component-wise approach: a simulation is accepted if and only if it satisfies all individual tolerance criteria simultaneously.

Here, $y_i \sim \pi(\cdot \mid \theta)$ corresponded to a single simulation from the underlying transmission process and $S^1(\cdot) \ldots S^L(\cdot)$ corresponded to the summary statistics calculated from a phylogeny.

1. Set the number of generations $T$, and the target number of particles $N$.

2. Initialise the tolerances $\varepsilon^1, \ldots, \varepsilon^X$. Set population indicator $t = 1$.

3. Set particle indicator $j = 1$.

4. If $t = 1$, sample $\theta^{**}$ independently from $\pi(\theta)$, otherwise, sample $\theta^*$ from the previous population $\{\theta_{t-1}\}$ with weights $\{W_t - 1\}$ and perturb the particle to obtain $\theta^{**} \sim Q_t(\cdot \mid \theta^*)$ according to a Gaussian random walk transition kernel $Q_t(\cdot)$.

5. If $\pi(\theta^{**}) = 0$, return to (4).

6. Simulate $i = 1, \ldots, n$ candidate data sets $y_i \sim \pi(\cdot \mid \theta^{**})$, and calculate:

$$\hat{\pi}(d \mid y) = \frac{\sum_{i=1}^{n} \mathbb{1}\left(|S^x(d) - S^x(y)| \leq \varepsilon_t^x,\ \forall x \in \{1, \ldots, X\}\right)}{n}. \tag{39}$$

7. If $\hat{\pi}(d \mid y) = 0$, then go to (4).

8. Set $\theta_t^{(j)} = \theta''$ and:

$$\text{if } t = 1, \quad W_t^{(j)} = \hat{\pi}(d \mid y), \tag{40}$$

$$\text{if } t > 1, \quad W_t^{(j)} = \frac{\hat{\pi}(d \mid y)\pi(\theta_t^{(j)})}{\sum_{j=1}^{N} W_{t-1}^{(j)} Q_t(\theta^{(j)} \mid \theta_{t-1}^{(j)})}. \tag{41}$$

9. If $j < N$, increment $j = j + 1$ and go to step 4).

10. Normalise the weights so that $\sum_{j=1}^{n_{part}} W_t^{(j)} = 1$.

11. If $t < T$, increment $t = t + 1$, initialise the tolerance $\varepsilon_{t+1}$ and go to 3).

We ran the ABC-SMC with eight simulations for each proposed $\theta$, 100 accepted particles per generation and 500 accepted particles for the last generation. Tolerance levels were chosen such that $\varepsilon_1^i \geq \ldots \geq \varepsilon_T^i \geq 0$, with $\varepsilon_{t+1}^i$ set to the 1/3 quantile of accepted distances at the end of iteration $t$. The algorithm terminated when $\varepsilon$ remained unchanged between generations or when the acceptance rate fell below 0.002 (i.e., more than 50,000 iterations required). If only one generation was completed, the procedure reduced to a simple rejection algorithm.

We assessed ABC-SMC convergence via the effective sample size of the normalised importance weights across all 48 transmission pairs using:

$$ESS_t = \frac{1}{\sum_{j=1}^{n}(W_t^{(j)})^2}. \tag{42}$$

where $W_t^{(j)}$ are the normalised particle weights at the final generation $t$. 41 of 48 pairs (85%) achieved adequate convergence (ESS >= 50; median 112, IQR 71-242). 3 pairs had lower ESS (20-50), yielding broader but still interpretable posteriors. 4 pairs had poor convergence (ESS < 20).

##### 1.2.6 Choice of summary statistics

From each inferred phylogeny, we computed five summary statistics: (1) topology class as defined by [13] (categorical: monophyletic–monophyletic, paraphyletic–monophyletic, and paraphyletic–polyphyletic); (2) root–tip consistency (categorical: consistent if the identity of the tip that minimises the number of internal nodes between itself and the root matches the documented direction of transmission; otherwise, inconsistent); (3) number of monophyletic groups from the exposed partner (integer); (4) phylogenetic diversity ratio (continuous: the sum of edge lengths in the transmitter's subtree divided by the sum of edge lengths in the exposed partner's subtree); and (5) tree imbalance measured by the Sackin index (continuous: the sum of ancestral nodes for each tip).

We verified whether the phylogenetic summary statistics could differentiate between four transmission scenarios arising from two factors: minimum versus maximum time since infection at transmission, and one versus two founding variants. For each transmission pair, we inferred phylogenies from 100 simulated genetic sequence sets per scenario, yielding 400 simulations per pair across the four scenarios.

The Sackin index and phylogenetic diversity ratio clearly distinguished between simulations assuming one versus two founding variants when the maximum reported TSI was used (39 of 43 pairs with uncertain TSI, or 91%), but were far less discriminatory when the minimum reported TSI was used (9 of 43, or 21%) (non-overlapping defined as separation of the 10th–90th percentile ranges on at least one axis; **Supplementary Figure 5**). This reduced power at minimum TSI likely reflects limited viral divergence: 24 of 43 pairs (56%) had a minimum TSI below six months. These phylogenetic measures also distinguished between minimum and maximum TSI scenarios in 19 of 43 pairs (44%).

The remaining three summary statistics (topology class, root-tip consistency, and number of exposed partner monophyletic groups) produced expected results. For example, MM and PM topologies (corresponding to one monophyletic group from the exposed partner) predominantly occurred with single founder variants, while PP topologies (corresponding to multiple monophyletic groups from the exposed partner) were more common with two founder variants (**Supplementary Figure 6**).

##### 1.2.7 Adequacy of accepted ABC-SMC simulations

The ABC-SMC fitting procedure accepted only simulations matching each transmission pair's empirical topology class and root-tip consistency. From these accepted simulations, we selected phylogenies whose number of monophyletic groups from the exposed partner, phylogenetic diversity ratio, and Sackin index best matched the empirical data. The median absolute differences between posterior medians and empirical values were 0 (IQR: 0–0) for the number of monophyletic groups from the exposed partner, 0.08 (IQR: 0.01–0.64) for the phylogenetic diversity ratio, and 16 (IQR: 1.0–62.8) for the Sackin index, indicating good overall agreement (**Supplementary Figure 7**). Examining whether posterior density distributions overlapped with the empirical values of the phylogenetic diversity ratio and Sackin index revealed exact matches for both metrics in 25 of 48 pairs (52%), for exactly one metric in 14 pairs (29%), and closest available matches in the remaining nine pairs (19%)

(**Supplementary Figure 8**).

#### 2 Supplementary Tables

Table 1: *

**Supplementary Table 1.** Model parameters and prior distributions.

| Fitted parameters | | | |
|---|---|---|---|
| | **Description** | **Prior Distribution** | **Model** |
| $p$ | Per-virion acquisition probability | $\sim$uniform(0, 1) | 1, 4, 5, 6, 7 |
| $p_E, p_A, p_L$ | Stage-specific per-virion acquisition probabilities | $\sim$uniform(0, 1) | 2, 3 |
| $f$ | Probability of permissive conditions for infection | $\sim$uniform(0, 1) | 1, 3, 5, 7 |
| $\tau_E$ | Duration of the early stage of infection (years) | $\sim$log-normal($\mu_{\log}$=-1.95, $\text{sd}_{\log}$=0.69) [3] | All |
| $c$ | Number of activated target cells available for initial infection in the exposed partner | $\sim$uniform(0, 1000) | 4, 5, 6, 7 |
| $r$ | Per-infected-cell establishment probability | $\sim$uniform(0, 1) | 4, 5 |
| $r_E, r_A, r_L$ | Stage-specific probabilities that an infected cell establishes systemic infection | $\sim$uniform(0, 1) | 6, 7 |
| $\varepsilon$ | Number of sexual exposures per year between an uninfected individual and an asymptomatically infected individual | $\sim$normal($\mu$=106.8, sd=23.5) [3] | All |

| Fixed parameters | | | |
|---|---|---|---|
| | **Description** | **Information** | **Reference** |
| $\tau_A$ | Duration of the asymptomatic stage of infection (years) | Varies with set-point viral load | [2, 14] |
| $\tau_L$ | Duration of the late stage of infection (years) | Ten months | [3] |
| $g_E(V)$ | Proportion of individuals with each viral load during the early stage | Combined viral load profiles from Fiebig stages I-V weighted by stage duration | [1] |
| $g_A(V)$ | Proportion of individuals with each viral load during the asymptomatic stage | Proportional to the relative lengths of the infectious periods of individuals with each SPVL | [2, 14] |
| $g_L(V)$ | Proportion of individuals with each viral load during the late stage | Given by the density of log viral load counts ($\mu$=5.1, sd=0.59) | [15] |
| $h(x, t)$ | Distribution of the number of genetic variants in a transmitter as a function of time since infection ($t$) | Main text | [14] |

Table 2: *

**Supplementary Table 2.** Target heterosexual empirical estimates for model calibration.

| **Data** | **Mean (SD)** | **MSM multiplier** | **Reference** |
|---|---|---|---|
| The relative (log) hazard of HIV acquisition during the early versus asymptomatic stage ($RH_E$) | 1.698 (1.084) | 1 | [3] |
| Per-act probability of HIV acquisition during the asymptomatic stage of infection | $7.2 \times 10^{-4}$ ($1 \times 10^{-4}$) | 4.16* | [16] |
| The relative hazard of HIV acquisition during the late versus asymptomatic stage ($RH_L$) | 5.0 (0.5†) | 1 | [3] |
| Per-act probability of multiple variant transmission | 0.187 (0.031) | 0.296 (0.043)‡ | [17] |
| The relative hazard of HIV multiple founders during the early versus asymptomatic stage ($RH_{MULT}$) | 3.125 (0.3**) | 1 | [4] |
| Annual Transmission probability per person by set-point viral load (measured in $\log_{10}$ copies/mL) during the asymptomatic stage of infection§ | 2.5: 0.019 (0.006)<br>3: 0.019 (0.006)<br>3.5: 0.074 (0.009)<br>4: 0.074 (0.007)<br>4.5: 0.143 (0.017)<br>5: 0.143 (0.013)<br>5.5: 0.143 (0.013)<br>6: 0.143 (0.013) | 1.57‖ | [18] |

*Average of receptive (1.38%) and insertive (0.11%) anal intercourse risks relative to heterosexual baseline (0.179%) [19], since per-act receptive anal risks do not differ between heterosexuals and MSM [20].

†Standard deviations assumed to represent uncertainty around reported point estimates.

‡Direct estimate (mean, SD) from [17] rather than a multiplier.

§Parameters derived by fitting normal distributions to the step function values from [18] (Figure 1A-Inferred relationships between SPVL and transmission rate), assuming the figure's lower, median, and upper estimates represent the 2.5th, 50th, and 97.5th percentiles.

‖Hazard of transmission for MSM vs heterosexuals, derived from US HIV incidence/prevalence rates for 2015 (CDC). Similar values observed for the US (1.39, 2014–2018, CDC) and UK (1.21, 2013–2022, UKHSA) using incidence/prevalence counts.

Table 4: *

**Supplementary Table 4.** Percentage distribution of single versus multiple virion and variant transmission by population and disease stage.

| | | Heterosexual population | | MSM |
|---|---|---|---|---|
| **Stage** | **Transmission scenario** | **Main** | **excluding $\mathbf{RH_{Mult}}^*$** | **population** |
| *Across all stages* | One virion and variant | 66 | 73 | 57 |
| | Multiple virions, One variant | 16 | 9 | 13 |
| | Multiple virions and variants† | 18 | 18 | 30 |
| *Early* | One virion and variant | 11 | 46 | 6 |
| | Multiple virions, One variant | 68 | 50 | 63 |
| | Multiple virions and variants | 21 | 4 | 31 |
| *Asymptomatic* | One virion and variant | 91 | 91 | 86 |
| | Multiple virions, One variant | 2 | 3 | 4 |
| | Multiple virions and variants | 7 | 6 | 10 |
| *Late* | One virion and variant | 63 | 60 | 39 |
| | Multiple virions, One variant | 3 | 4 | 3 |
| | Multiple virions and variants | 34 | 36 | 58 |

*The relative hazard of HIV multiple founders during the early versus asymptomatic stage
†Corresponds to the fitted probability of multiple variants founding infections. Empirical observations used for model fitting: 18.6% for heterosexual populations and 29.6% for MSM.

Table 5: *

**Supplementary Table 5.** Model predictions across all viral loads and infection stages.

| **Estimate** | **Mean (median, 95% HPD)** | | |
|---|---|---|---|
| | **Heterosexual population** | | **MSM population** |
| | **Main** | **Excluding $\mathbf{RH_{Mult}}$** | |
| Probability of acquisition per sex act | 0.12%(0.12, 0.09-0.15) | 0.11%(0.10, 0.08-0.20) | 0.40%(0.39, 0.25-0.56) |
| Expected proportion of target cells infected, given permissive conditions | 0.42 (0.42, 0.35-0.47) | 0.40 (0.41, 0.29-0.47) | 0.41 (0.41, 0.35-0.46) |
| Probability of transient infection, given cells infected | 0.82 (0.82, 0.77-0.86) | 0.82 (0.83, 0.68-0.93) | 0.76 (0.76, 0.70-0.81) |
| Probability of transient infection per sex act | 0.53%(0.51, 0.33-0.82) | 0.65%(0.49, 0.20-2.62) | 1.24%(1.21, 0.70-2.04) |
| Ratio of transient to established infections | 4.6 (4.5, 3.3-6.3) | 5.2 (4.7, 2.0-13.2) | 3.1 (3.1, 2.3-4.2) |

### 3 Supplementary Figures

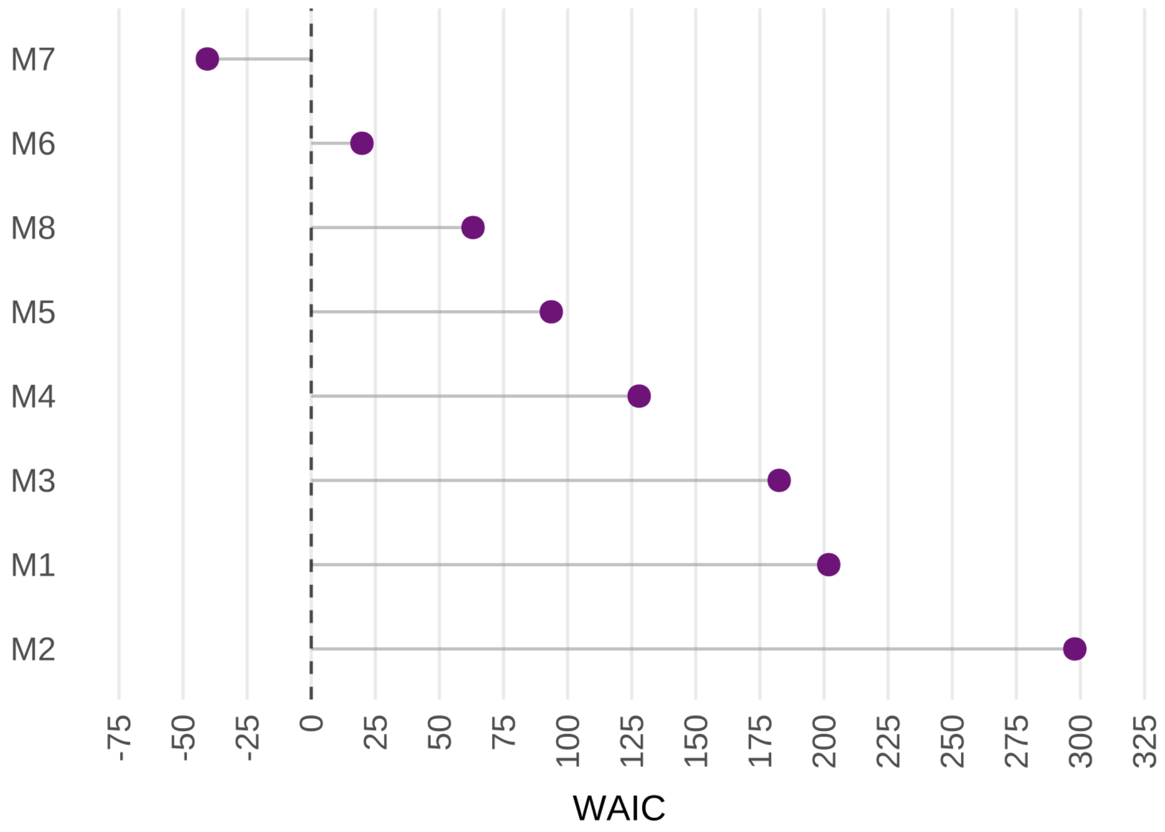

Figure 1: *

**Supplementary Figure 1. Model M7 achieves the lowest Watanabe–Akaike information criterion (WAIC) among all seven models.** Lower WAIC values indicate better model fit. Seven distinct models are compared using different colours: M1 (Intermittent susceptibility), M2 (Stage-dependent virus infectivity), M3 (Intermittent susceptibility, stage-dependent virus infectivity), M4 (target-cell-limited), M5 (Intermittent susceptibility & target-cell-limited), M6 (stage-dependent virus infectivity & target-cell-limited), and M7-M8 (Intermittent susceptibility, stage-dependent virus infectivity and target-cell-limited). M7 and M8 differ in that M7 assigns stage-specific differences to per-infected-cell establishment probabilities , whereas M8 assigns them to per-virion acquisition probabilities.

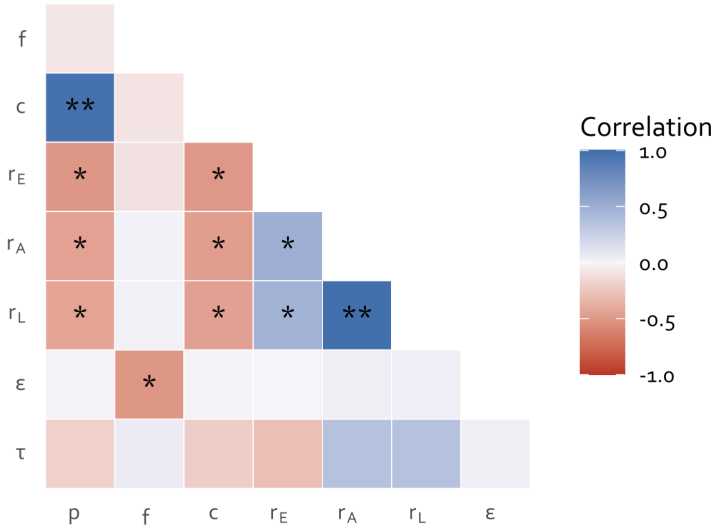

Figure 2: *

**Supplementary Figure 2. Fitted parameters of M7 show strong correlations reflecting a trade-off between cell infection and establishment.** Asterisks indicate strong and statistically significant correlations ($p < 0.001$): $|r| \geq 0.9$ (**) and $|r| \geq 0.4$ (*).

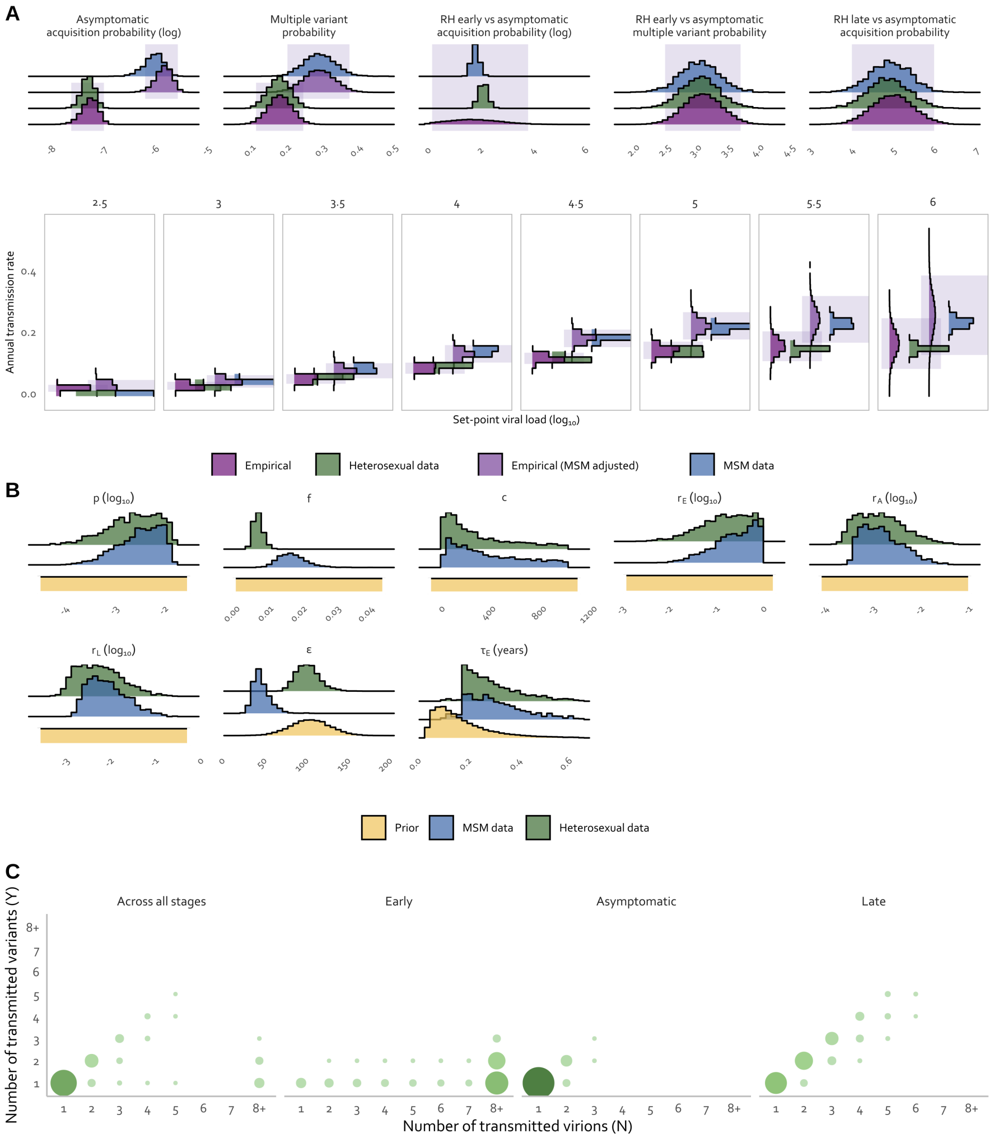

Figure 3: *

**Supplementary Figure 3. The model recalibrated to MSM data fits all six observations and predicts a higher permissive probability. (A)** MCMC fit to assumed empirical distributions. **(B)** Parameter posterior estimates. **(C)** Joint probability of the effective number of transmitted virions and variants founding infection in the exposed partner (Greater than or equal to 0.005)

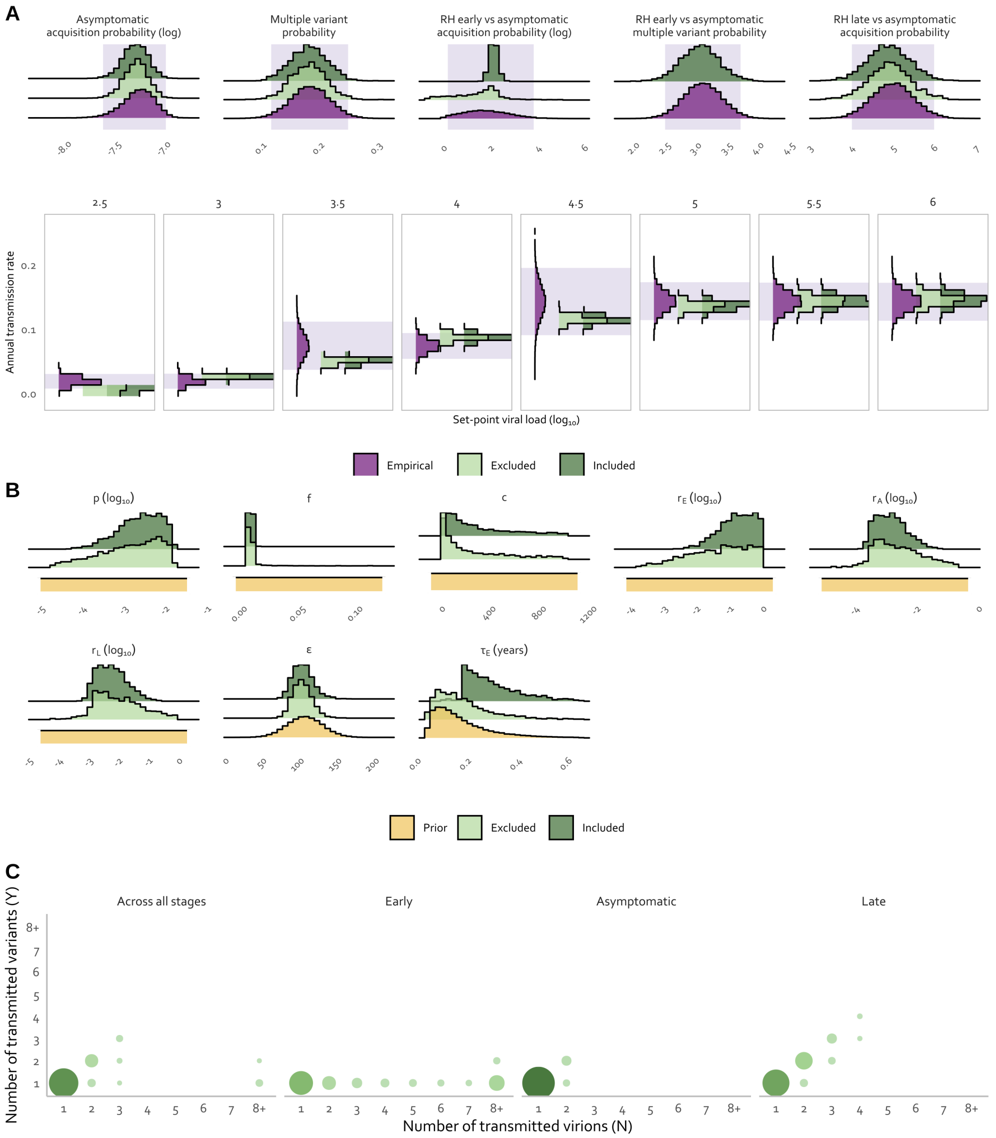

Figure 4: *

**Supplementary Figure 4. Excluding $\mathbf{RH_{Mult}}$ broadens credible intervals for stage-specific establishment probabilities. (A)** MCMC fit to the heterosexual population data. **(B)** Parameter posterior estimates. **(C)** Joint probability of the effective number of transmitted virions and variants founding infection in the exposed partner (Greater than or equal to 0.005)

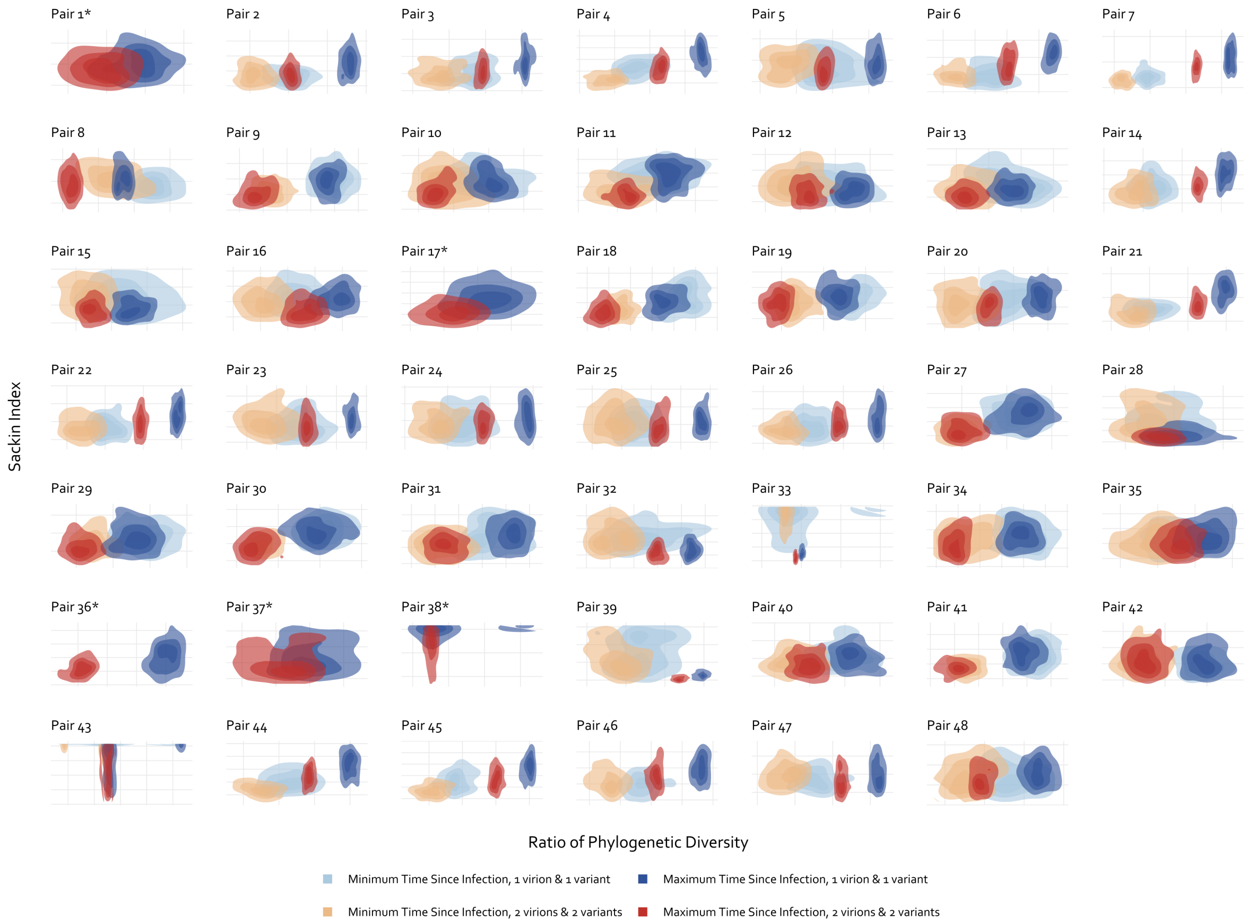

Figure 5: *

**Supplementary Figure 5. The Sackin index and phylogenetic diversity ratio distinguish variant scenarios at maximum but not minimum TSI.** Phylogenetic measures were derived from trees of simulated nucleotide sequences using linked transmission-pair data. Colours represent test scenarios combining the transmitter's infection duration (minimum or maximum) and transmitted variant scenarios (one virion/one variant or two virions/two variants). Asterisk (*) indicates pairs with uncertainty in the viral load at transmission only. For these pairs, only the single- and multiple-variant scenarios were evaluated and are presented using the maximum time since infection colour scheme (dark colours).

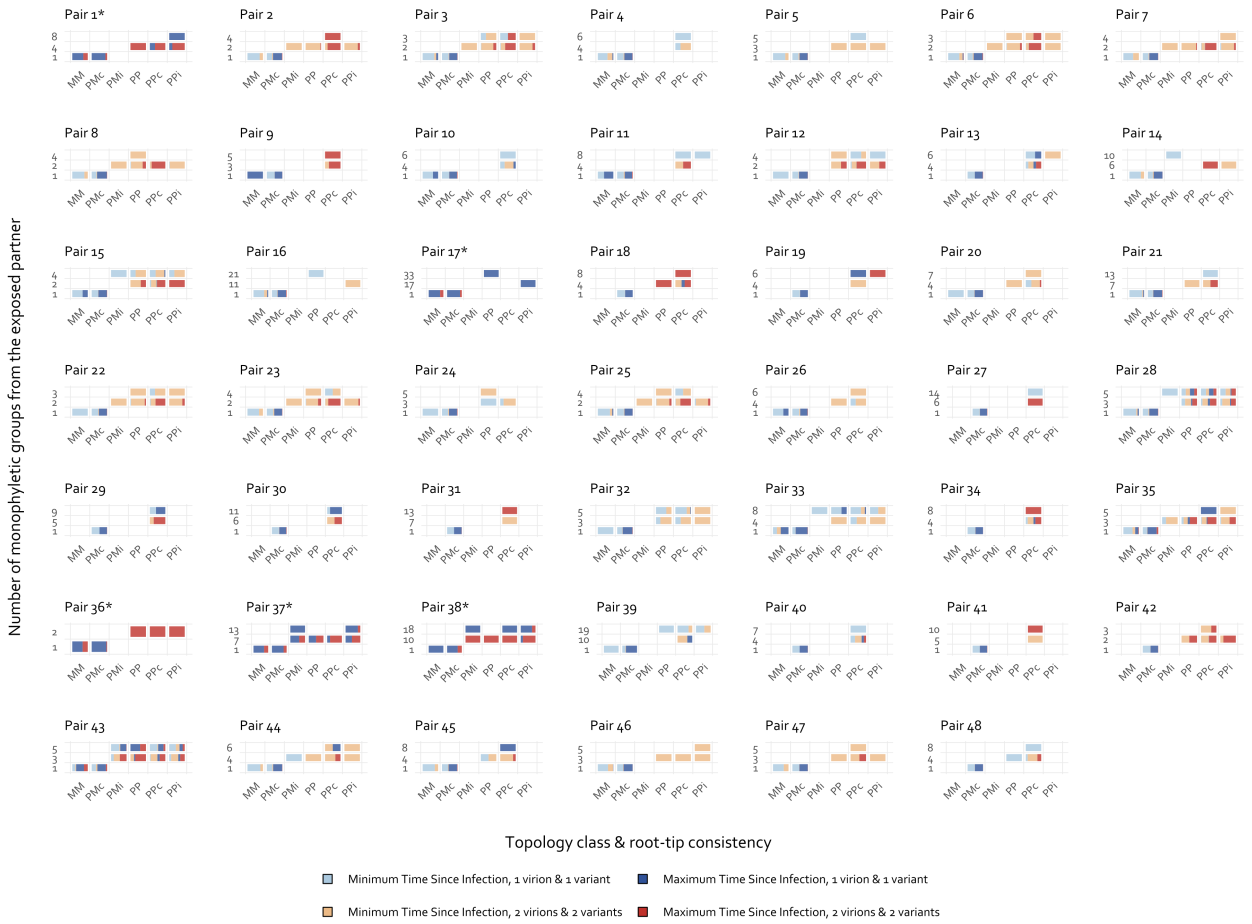

Figure 6: *

**Supplementary Figure 6. Topology class and monophyletic group counts show expected but less discriminatory patterns across scenarios.** Phylogenetic measures were derived from trees of simulated nucleotide sequences using linked transmission-pair data. Colours represent transmission scenarios combining the transmitter's infection duration (minimum or maximum) and transmitted variant scenarios (one virion/one variant or two virions/two variants). Asterisk (*) indicates pairs with uncertainty in the viral load at transmission only. Each bar represents the proportion of simulations from a given transmission scenario. For simplicity, monophyletic groups from the exposed partner are presented for one, two, mid and maximum values only. For topology class: Monophyletic-monophyletic (MM), Paraphyletic-monophyletic (PM), or Paraphyletic-polyphyletic (PP). For root-tip consistency: consistent ($PM_c$, $PP_c$) if the identity of the tips that minimises the number of internal nodes between itself and the root matches the documented direction of transmission; otherwise, inconsistent ($PM_i$, $PP_i$) or indeterminate (MM, PP).

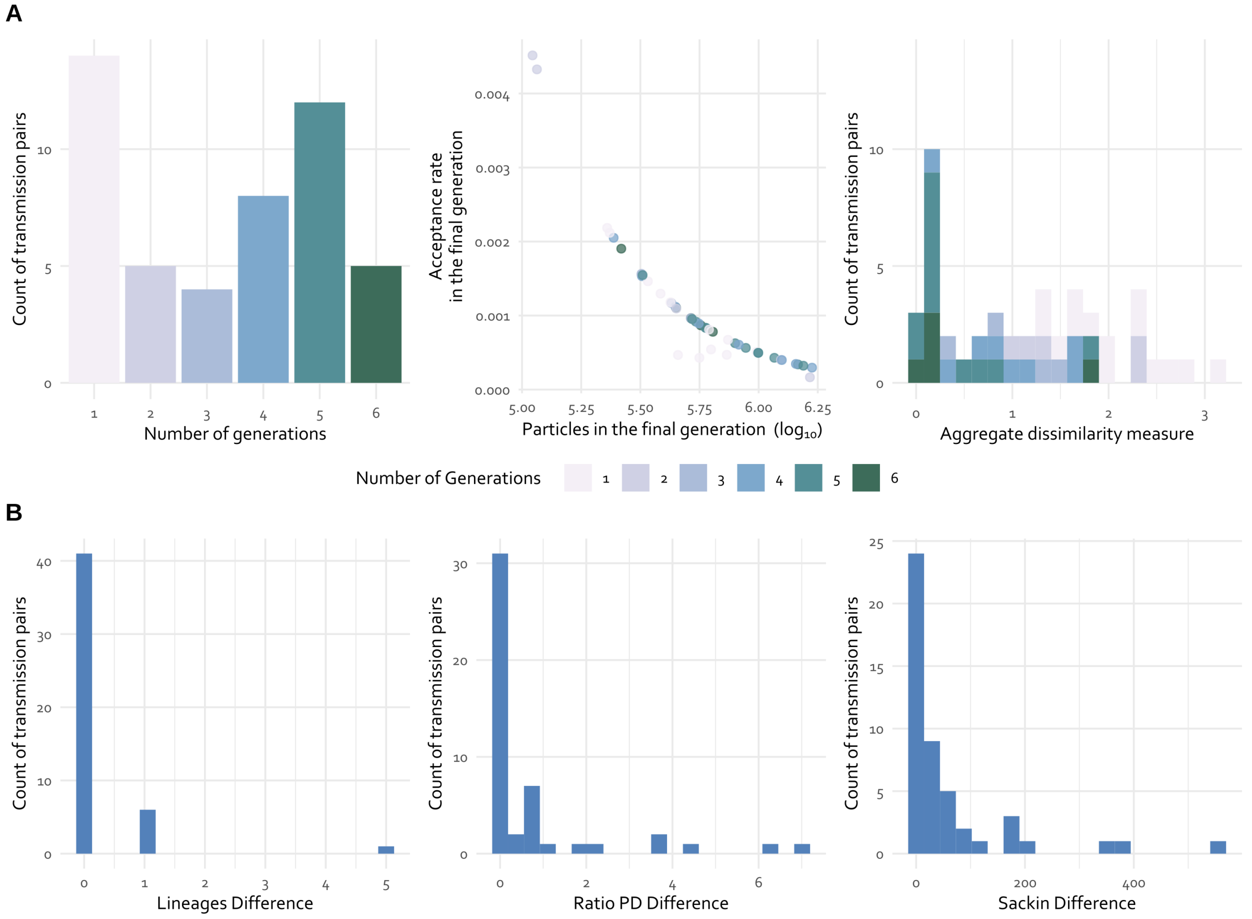

Figure 7: *

**Supplementary Figure 7. The ABC-SMC procedure converged for most transmission pairs with low median distances between simulated and empirical data. (A)** Distribution of the number of generations, the acceptance rate and the number of particles in the final generation. To quantify similarity between simulated and empirical data, we calculated a composite distance measure. For each tree pair, we computed absolute differences between summary statistics, using log transformation for quantitative variables and binary comparisons for qualitative variables. The final distance was the mean across all statistics, where values closer to 0 indicate a better fit. **(B)** Distribution of the median differences between simulated and empirical data across accepted particles in the final ABC generation for each transmission pair. Topology class and root-tip consistency are not shown as accepted simulations were constrained to match the empirical values for these statistics.

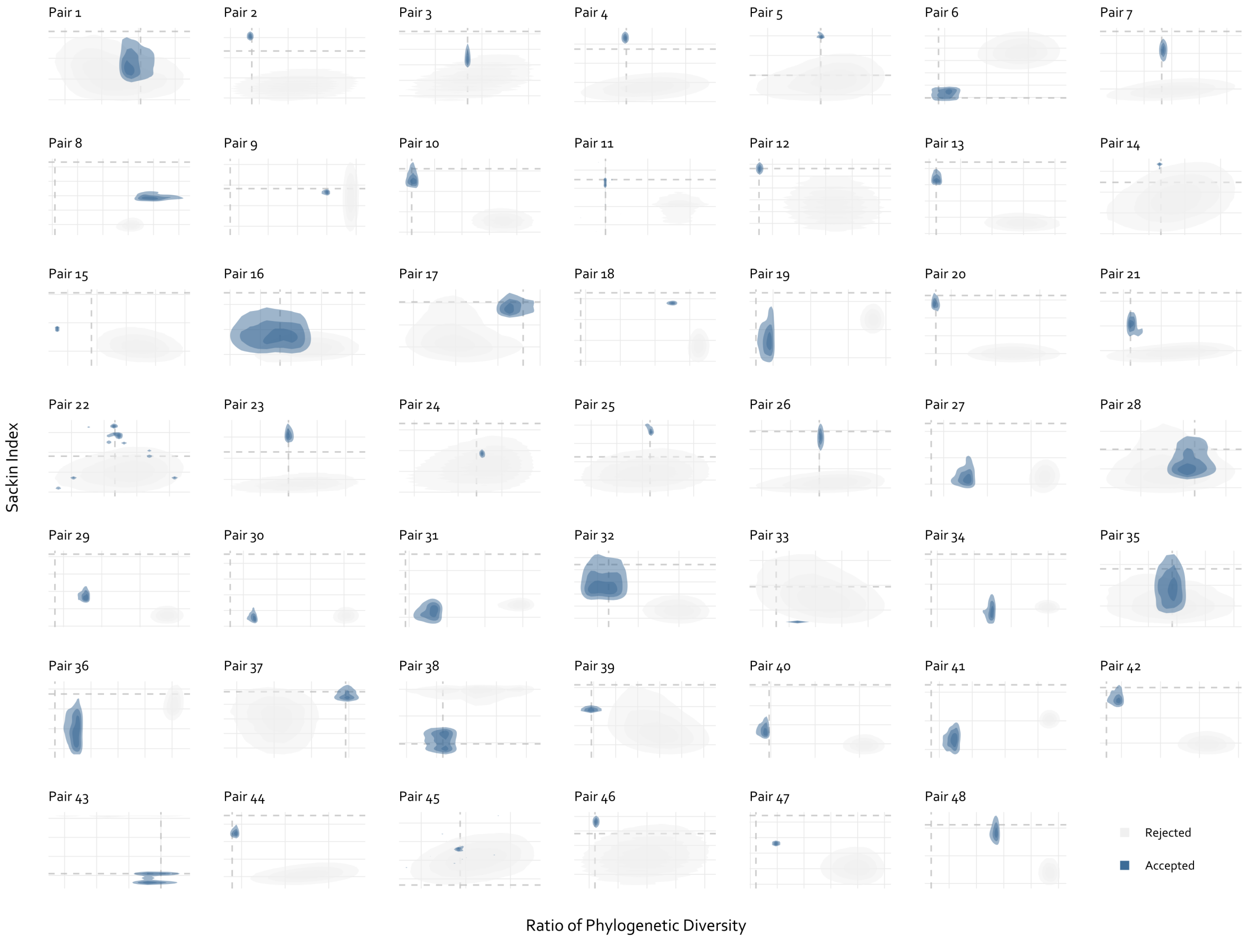

Figure 8: *

**Supplementary Figure 8. Accepted ABC-SMC particles cluster near empirical phylogenetic diversity and Sackin index values for most pairs.** The fit is shown only for the final generation. Accepted and rejected particles are shown as blue and light grey, respectively. Dashed lines intersect at the Sackin index and phylogenetic diversity ratio derived from empirical data, with point proximity indicating similarity between simulated and empirical phylogenies.

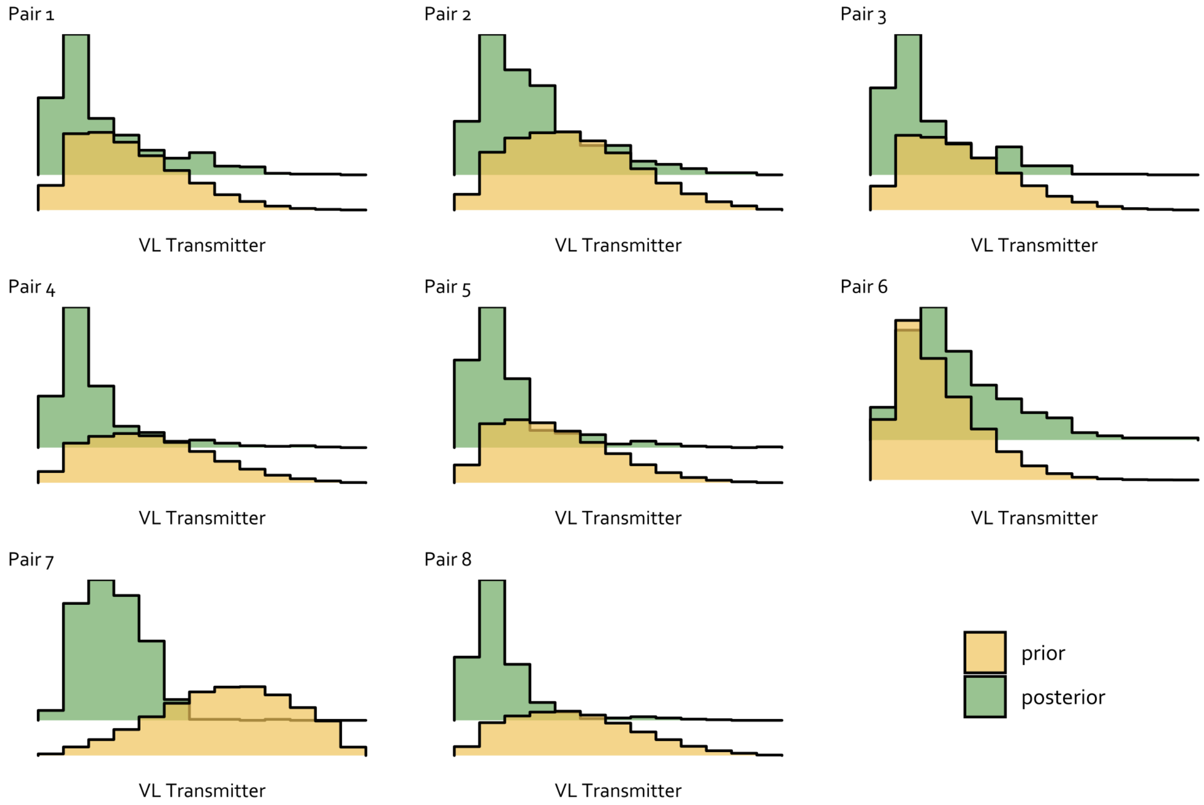

Figure 9: *

**Supplementary Figure 9. Phylogenetic data narrowed viral load posteriors for the majority of early transmission pairs.** Each panel represents one transmission pair. Prior distributions reflect a clinically plausible range of $\log_{10}$ viral load, and posterior distributions incorporate phylogenetic data through an ABC-SMC fitting procedure.
